# Population-scale plasma proteomics reveals pre-diagnostic biomarkers and shared pathways of cancer risk across 25 cancers

**DOI:** 10.64898/2026.09.22.26363676

**Authors:** James Yarmolinsky, Vivian Viallon, Vernon A Burk, David C Muller, P Martijn Kolijn, Christina M Lill, Kostas Tsilidis, Pietro Ferrari, George Richenberg, Ziqiao Wang, María Dolores Chirlaque López, Ana Jiménez-Zabala, Rosario Tumino, Carlotta Sacerdote, Nicholas Wareham, Zhe Huang, Karl Smith-Byrne, Ruth Travis, Nilanjan Chatterjee, Elizabeth A Platz, Roel CH Vermeulen, Elio Riboli, Marc J Gunter

## Abstract

The causes of most cancers remain unknown because the biological pathways underlying cancer development are poorly understood. Large-scale plasma proteomics offers a powerful strategy to uncover these pathways, identify biomarkers for cancer early detection, and discover novel therapeutic targets but has rarely been applied in prospective cohorts. Here, we profiled 6,402 plasma proteins using the SomaScan^TM^ 7K Assay in baseline samples from 9,828 participants in the European Prospective Investigation into Cancer and Nutrition (EPIC), including 6,073 incident cancer cases within a case-cohort design. We identified 1,732 protein-cancer associations (FDR *P* < 0.05) across 25 cancer endpoints in multivariable-adjusted models, including 60 proteins associated with overall cancer risk. Confirmation analyses in the Atherosclerosis Risk in Communities (ARIC) Study cohort showed directional concordance for 78% of associations and statistical replication of 49% of findings. Most proteins were associated with a single cancer type, whereas 284 were associated with multiple cancers, indicating both shared and distinct biological pathways across cancer sites. Stratification by follow-up time revealed 523 protein associations that were restricted to specific time windows after blood collection (<2 years, <5 years, and >5 years), including 447 that were not apparent in primary analyses, highlighting proteins with potential roles as early detection or aetiological biomarkers. Proteins associated with cancer risk were enriched across diverse biological pathways and 315 corresponded to targets of approved cancer and non-cancer medications, while most proteins represent previously unrecognised links to cancer risk. As the most comprehensive prospective investigation of the plasma proteome and cancer risk to date, our study significantly expands our understanding of key biological pathways shaping cancer development and provides a resource for the scientific community to accelerate development of early detection strategies, biomarkers for risk stratification, and novel therapeutic interventions for cancer prevention.

## Background

The global burden of cancer is increasing, driven by population ageing and growth along with changes in exposure to risk factors(*1, 2*). Though advances in treatment have led to notable improvements in cancer survival in recent decades, robust cancer prevention and early detection strategies remain critical tools for reducing the population cancer burden(*3, 4*). Developing these strategies requires deeper understanding of the biological processes that precede cancer diagnosis and the identification of biomarkers that can enable earlier detection and inform preventive interventions. Yet pre-diagnostic biology has until recently been inaccessible at population-scale, limiting the development of cancer predictive markers, early detection assays, and preventive therapies that would meaningfully reduce incidence and mortality from this disease.

Recent advances in high-throughput proteomic profiling using affinity-based technologies now enable the measurement of thousands of plasma proteins simultaneously from a single biospecimen(*5, 6*). Plasma proteins represent a particularly informative molecular layer for studying cancer development as many proteins are actively secreted or released into the circulation during physiological and pathological processes, including tumour growth, immune activation, metabolic dysregulation, and tissue damage(*7–9*). Indeed, several widely used clinical biomarkers for cancer detection and monitoring are circulating proteins, including prostate-specific antigen (PSA) for prostate cancer, cancer antigen-125 (CA-125/MUC16) for ovarian cancer, and alpha-fetoprotein (AFP) for hepatocellular carcinoma(*10–12*). The integration of proteomic platforms into large-scale population-based studies can thus provide previously unparalleled insights into key biological pathways underlying tumour development, accelerating the development of novel early detection, risk stratification, and therapeutic prevention strategies for cancer.

To date, prospective proteomic studies have identified numerous protein-cancer associations, but have been largely constrained by limited sample sizes, restricted proteome coverage, and/or a focus on a small number of cancer types(*13–22*). As a result, the extent to which circulating protein signatures are cancer-specific, shared across tumour types, or vary across stages of disease development remains unclear.

Here, we performed plasma proteomic profiling of 6,402 proteins using the SomaScan 7K Assay in baseline samples from 9,828 participants in the European Prospective Investigation into Cancer and Nutrition (EPIC), including 6,073 incident cancer cases within a case-cohort design. This scale enabled a comprehensive evaluation of associations between circulating proteins and risk across 25 distinct cancer endpoints. We systematically assessed protein-cancer associations and evaluated the robustness of findings through confirmation analyses in an independent prospective cohort. We then examined shared and cancer-specific proteomic signatures, evaluated associations according to time from blood collection, characterised implicated biological pathways, and explored opportunities for drug repurposing for cancer prevention by mapping proteins to approved therapeutic targets.

## Materials and Methods

### The EPIC multi-endpoint case-cohort study

EPIC is a prospective cohort comprising >520,000 individuals recruited in 1992-2000 from 23 centres across 10 European countries (Denmark, France, Germany, Greece, Italy, Norway, Spain, Sweden, the Netherlands, and the UK)(*23*). At recruitment, individuals were between 35-70 years of age, ∼70% were women, and non-fasting citrate plasma samples were collected from ∼75%. A multi-endpoint case-cohort study was established within EPIC, whereby plasma samples from a total of 17,841 individuals from the UK, the Netherlands, Spain, and Italy underwent proteomic analysis using the SomaScan 7K Assay. Our study focused on data from 9,828 individuals from the EPIC case-cohort, namely (i) the subcohort component, a random subsample of the eligible EPIC participants comprising 4,115 participants, including 360 incident cancer cases, and (ii) the cancer component which comprised 5,713 additional incident cancer cases randomly selected among the eligible EPIC participants who developed one of the most frequent cancer types during follow-up (bladder, breast, colorectal, endometrial, oesophageal, glioma, kidney, liver, lung, leukemia, lymphoma, melanoma, multiple myeloma, ovarian, pancreas, prostate, stomach, thyroid, and upper aero-digestive tract).

### Cancer endpoints

In EPIC, incident first primary cancer cases (excluding non-melanoma skin cancers) were identified through a combination of centre-specific methods, including health insurance records, cancer and pathology registries, and active follow-up through study participants and their next-of-kin. Follow-up for each participant and event of interest began upon inclusion in the study and ended upon the occurrence of the event, loss to follow-up, or the last date of ascertainment, whichever came first. In the present study, cancer endpoints were defined as the first incident cancer diagnosis, coded using the 10th revision of the World Health Organisation’s International Statistical Classification of Diseases (ICD-10). We considered overall cancer as well as 24 specific cancer subtypes, namely upper aero-digestive tract (C00–14, C32, including two subtypes: squamous cell, and others), oesophagus (C15; including two subtypes: squamous cell, and others), stomach (C16), colon (C18) and rectum (C19–20), liver (C22; including two subtypes: hepato-cellular carcinoma and intra-hepatic cancer, and extra-hepatic and gallbladder cancer), pancreas (C25), lung (C34), malignant melanoma (C43), breast in women (C50, including two subtypes: pre- and post-menopausal), endometrial (C54), ovary (C56), prostate (C61), kidney (C64–65), bladder (C67), brain/glioma (C71), thyroid (C73), and three subtypes of blood cancers: non-Hodgkin lymphoma (NHL; C82–85), multiple myeloma (C90), and chronic lymphocytic leukemia (C91.1).

### Proteomics data: the SomaScan 7K Assay, pre-processing and quality control

Assays were performed using SomaScan reagents (aptamers) according to the manufacturer’s detailed protocol(*24*). The SomaScan platform uses modified nucleotides (Slow Off-rate Modified Aptamers-SOMAmers^TM^) which specifically bind to protein targets and quantifies them in relative fluorescence units (RFUs) using DNA microarray. Separate SOMAmers can bind to different epitopes on the same protein (which can be influenced by post-translational modifications or protein complexes), enabling a larger number of proteins and protein complexes to be quantified.

In our main analysis, we used RFUs normalized by Somalogic through the following steps: hybridization normalization, intraplate median normalization, plate scaling and calibration, and adaptive normalization to a population reference. The normalized RFUs were then log-transformed to reduce skewness. Samples for which any normalization scale factor was outside [0.4-2.5] and those detected as outliers using an approach based on PCA and the local outlier factor statistic were excluded. Leveraging blind duplicates (n=233) generated from the EPIC study, intra-class correlation coefficients were computed to assess reproducibility of the SOMAScan assay. These preliminary analyses (see **Supplementary Methods**, **fig. S1**) suggested that plate correction could improve reproducibility for some SOMAmers, which was then performed using a residual approach: for each SOMAmer, its measurements were corrected for plate effect, with plate effect estimated in linear mixed effect models adjusted for centre, age, sex, body mass index, smoking status, and incidence of cancer, cardiovascular disease, type 2 diabetes, neurodegenerative diseases, and death, to preserve possible biological variation due to these factors. Finally, measurements of each SOMAmer were centred and scaled so that their mean and standard deviation were respectively 0 and 1 in the subcohort.

Several alternative versions of the pre-processing strategy described above were further considered in sensitivity analyses. These included versions without plate correction or without adaptive normalization to a population reference, versions in which log-transformed relative abundances were capped at greater or less than 5 standard deviations from the mean, and versions using inverse-rank normalization instead of log-transformation. Overall, the choice of pre-processing version did not substantially affect the results (**fig. S2-3**).

### Statistical analyses

All analyses were conducted using R version 4.1.2. Associations between 7,289 SOMAmers and 25 cancer endpoints (risk of 24 individual cancer sites and overall cancer) were studied in Cox proportional hazards models using age as the underlying time scale. Prentice weights and robust variance were used to account for the case-cohort design. In particular, for each individual cancer site, analyses included the 4,115 subcohort members plus the incident cases of that specific cancer site, while incident cases of other cancer sites outside the subcohort were ignored. Minimally adjusted models were stratified on sex, centre, and age at recruitment (5-year categories). Multivariable-adjusted models were additionally adjusted for cancer-specific risk factors to facilitate the identification of novel cancer-specific risk factors, as detailed in the **Supplementary Methods**. Analyses were also performed in men and women separately. We used a FDR (false discovery rate)-adjusted *P*-value (termed “*q*-val”) threshold of 0.05 to identify evidence for statistical associations, with FDR accounting for the K*7,289 statistical tests, with K the number of considered cancer endpoints (i.e., K=25 for all analyses except for analyses restricted to the first two years of follow-up where K=5, representing overall cancer and the 4 most common cancer sites evaluated, and, male-specific analyses where K=21, and female-specific analyses where K=24). To assess heterogeneity of aptamer associations across cancer types, we fitted Prentice-weighted Cox proportional hazards models with cancer type-specific interaction terms. Because likelihood ratio tests are not appropriate with the robust variance estimator used for Prentice-weighted case-cohort analyses, heterogeneity was evaluated using a multivariable Wald test of the null hypothesis that all interaction coefficients were equal to zero, based on the robust sandwich covariance matrix. All statistical tests were two-sided.

Confirmation analyses for FDR-significant EPIC aptamer-cancer associations were performed in the Atherosclerosis Risk in Communities (ARIC) study for the 4,712 proteins measured in both studies(*25, 26*). ARIC is a prospective cohort study of 15,972 participants recruited from Forsyth County, North Carolina; Jackson, Mississippi; suburbs of Minneapolis, Minnesota; and Washington County, Maryland in the US. 3,342 incident cancers were diagnosed over a median 22.6 years of follow-up. 4,712 unique plasma proteins (4,955 aptamers) were measured in EDTA plasma samples at Visit 2 from 9,391 participants using the SomaScan 5K assay. Participants contributed person-time from Visit 2 to their first cancer, death, or 12/31/2015. We tested all FDR-significant aptamer-cancer associations identified in EPIC where there was 80% statistical power in ARIC to detect a hazard ratio (HR) equivalent to or more extreme than that in EPIC. Using the prospective cohort design, HRs and 95% confidence intervals (CIs) for incidence of each cancer site were tested using Cox proportional hazards regression models with covariate adjustment strategies comparable to those employed in EPIC where possible. ARIC and EPIC differ in participant characteristics (i.e. 76% of participants in ARIC were of European ancestry and 24% were of African ancestry, whereas in EPIC >98% of participants were of European ancestry), median follow-up time (22.6 years in ARIC vs 13.1 years in EPIC), and plasma anticoagulant used for sample collection (EDTA vs citrate). Further information on statistical analyses is presented in the **Supplementary Materials**.

### Follow-up time stratified analyses

We examined whether there was evidence of an association when analyses were restricted to different follow-up periods (i.e. <2 years, <5 years, and >5 years since blood collection). Where there was evidence (*q*-val < 0.05) of an association of a protein with a cancer outcome within any of these follow-up periods, we further verified that findings were specific to the follow-up period by using time-dependent coefficient Cox models to model time-varying effects across the full follow-up duration and by visually inspecting the shape of the association of time-varying hazard ratios with time since blood collection. We restricted analyses with <2 years of follow-up to overall and to common cancers (post-menopausal breast cancer, colon cancer, lung cancer, prostate cancer) given the smaller number of cancer cases diagnosed within this follow-up period. We used “minimally adjusted” models (i.e. stratified on sex and centre and adjusted for age at recruitment) for analyses restricted to <2 years and <5 years of follow-up and a “multivariable-adjusted” model (i.e. further adjustment for known cancer type-specific risk factors) for analyses restricted to >5 years of follow-up. The rationale for using minimally-adjusted models in analyses restricted to the first 2 years and 5 years of follow-up and multivariable-adjusted models in >5 year follow-up stratified analyses is that proteins identified in the former analyses are more likely to represent non-causal biomarkers of latent or early-stage cancer (e.g. biomarkers for early detection) whereas the latter are more likely to reflect proteins with possible aetiological roles in cancer risk, necessitating comprehensive confounder adjustment. Follow-up time stratified analyses were performed separately for aptamer-cancer associations that were *q*-val < 0.05 and *q*-val ≥ 0.05 in primary analyses.

### Over-representation analysis, tissue-specificity, functional annotation of proteins, and drug repurposing evaluation

To examine tissue-specific expression patterns of cognate genes for proteins associated with cancer risk, we performed MAGMA (Multi-marker Analysis of GenoMic Annotation) tissue specificity analysis as implemented in FUMA (Functional Mapping and Annotation of GWAS)(*27*). We also provided biological characterisation of proteins by performing over-representation analysis by comparing all FDR-significant proteins to all SomaScan 7K proteins evaluated using Reactome, GO (Gene Ontology), and Human MSigDB (Molecular Signatures Database) Collections Hallmark gene sets as implemented in clusterProfiler(*28–31*). The Human Protein Atlas (HPA) was used to functionally annotate proteins as intracellular, secreted, or membrane-bound. We annotated proteins to pharmacological targets of approved drugs with reference to DrugBank and ChEMBL(*32, 33*).

A schematic overview of the study design is presented in **Fig. 1**.

**Fig. 1:**
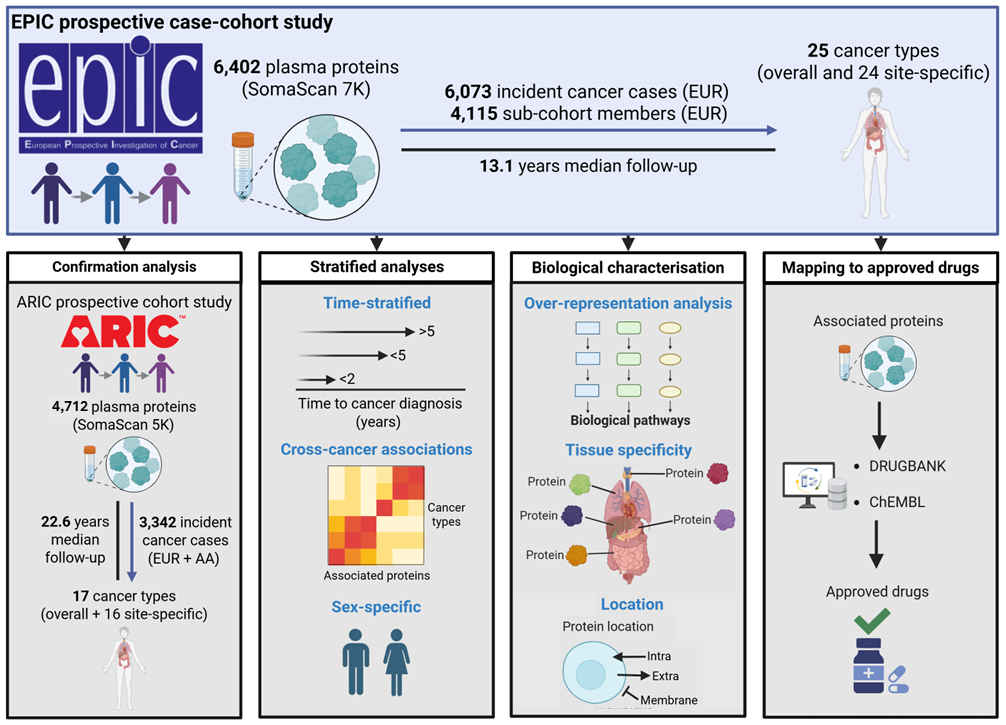
Schematic overview of the study design.

## Results

The EPIC case-cohort included 6,073 incident cancer cases (including 360 from the subcohort and 5,713 outside the subcohort) and 3,755 participants of the subcohort who did not develop cancer, with a median 13.1 years (Q1:8.1, Q3:15.8) follow-up. The number of incident cases inside and outside the subcohort, and the number of “controls” in the subcohort is presented in **table S1** for each cancer type included in this analysis overall and stratified by sex. Baseline characteristics of the EPIC case-cohort are provided in **table S2**. Compared to sub-cohort members, individuals who developed cancer were more likely to be men, current or former smokers, and to have higher alcohol intake.

### Association of plasma proteins with cancer risk

Across 25 cancer endpoints, we identified 1,732 protein–cancer associations (1,820 aptamer–cancer associations) at *q*-val < 0.05 in multivariable-adjusted models, including 60 proteins associated with overall cancer risk. An overview of the strongest associations between aptamers and each cancer type is shown in **Fig. 2**, with complete FDR-significant results provided in **tables S3-27**. Multivariable adjustment led to a median 37% (Q1:18%, Q3:60%) attenuation of the number of FDR-significant associations across cancer sites suggesting important roles of hypothesised risk factors in confounding associations in minimally adjusted models (**Fig. 3, table S28**).

**Fig. 2:**
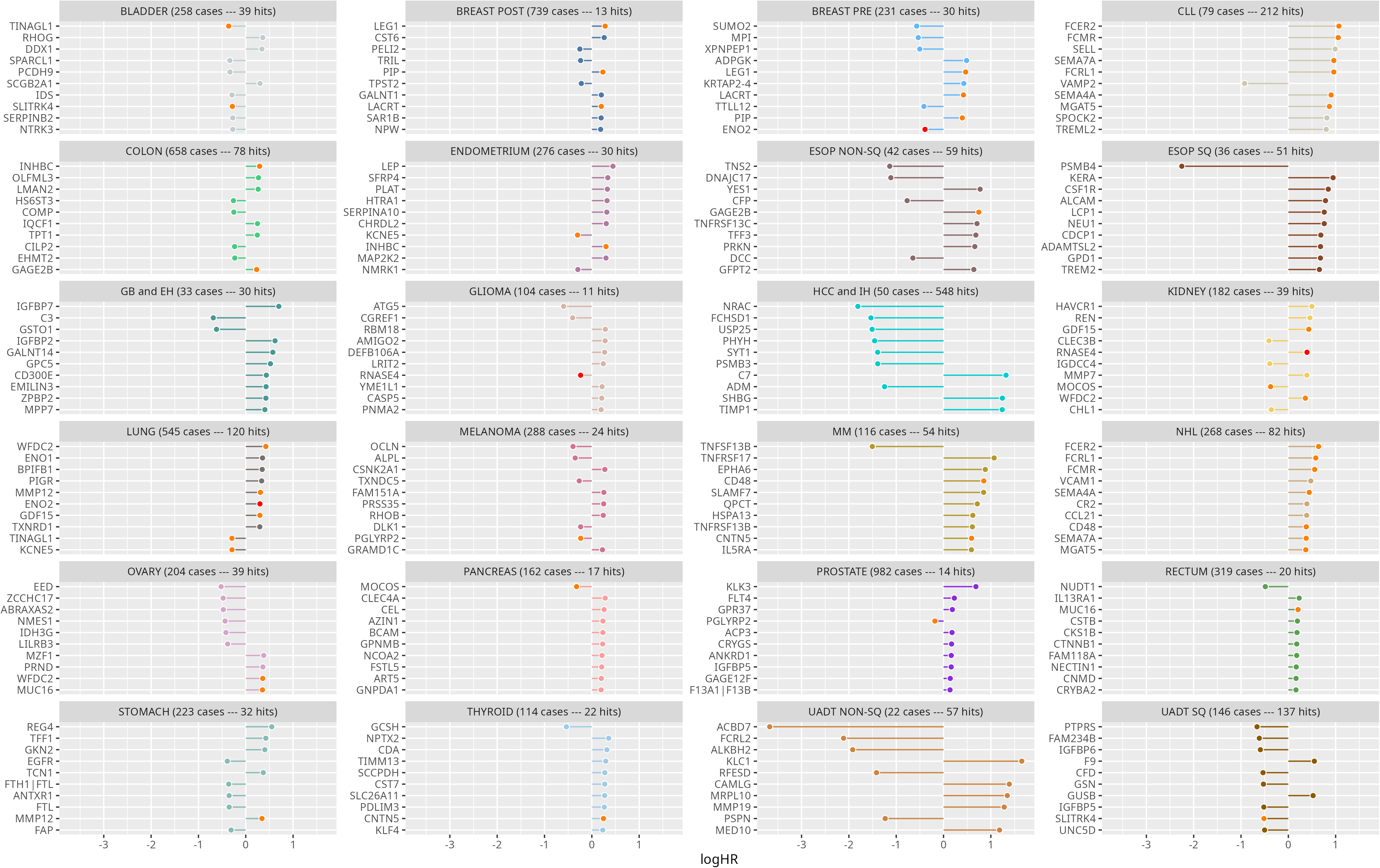
Overview of the strongest associations between aptamers and each cancer type by magnitude of logHR.

**Fig. 3:**
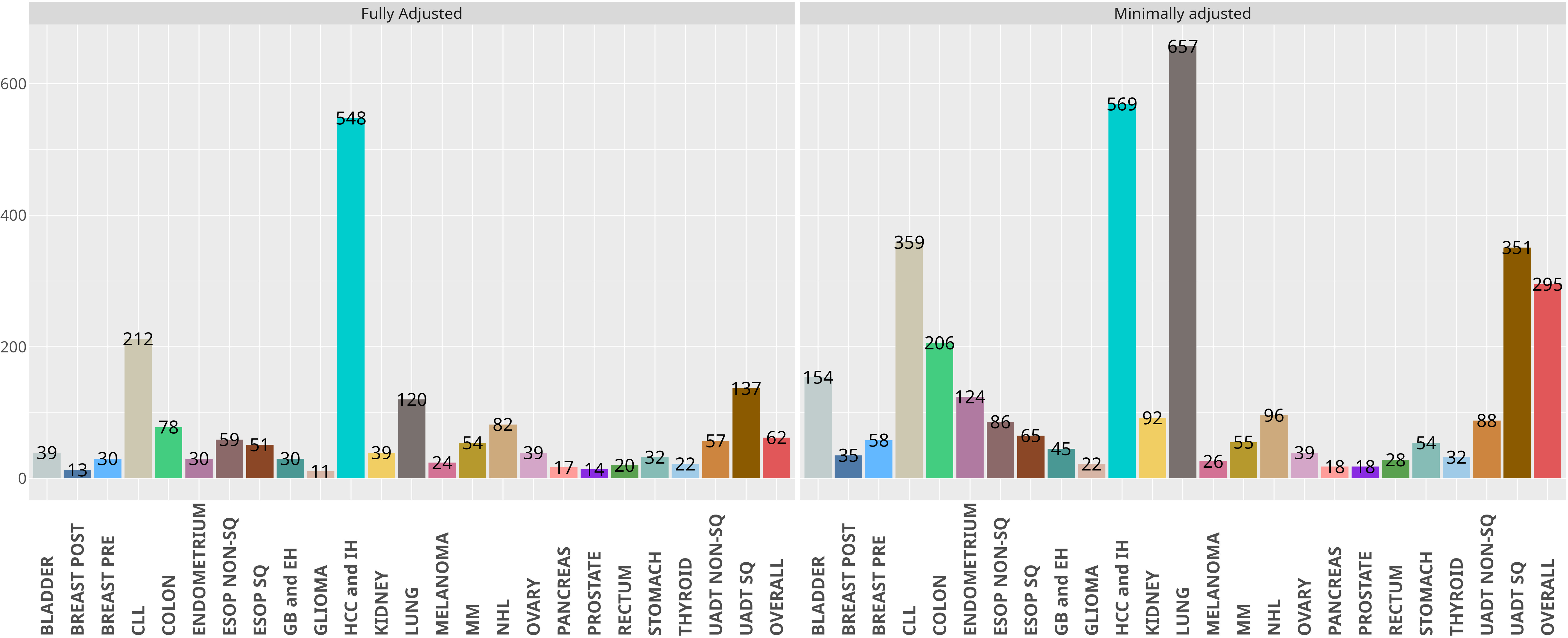
Comparison of number of FDR-significant aptamer-cancer associations per cancer type across minimally and fully adjusted models.

Most proteins (79.3%, 1,047/1,320) were associated with only one cancer type with 13 associated with 4 or more cancer endpoints, including MUC16, TNFRSF1B, and PIGR. The number of protein associations per cancer type varied markedly, ranging from 502 for hepatocellular carcinoma and intrahepatic bile duct carcinoma (IH) and 204 for chronic lymphocytic leukemia (CLL) to 13 for post-menopausal breast cancer and 11 for both prostate cancer and glioma (median 37 proteins per cancer type, Q1:23.5, Q3:63). Around 30% (502/1,672) of all cancer type-specific associations were observed for hepatocellular carcinoma despite the relatively low number of cases for this cancer type (N=50), likely reflecting the central role of the liver in protein synthesis and secretion along with the potential enrichment for protein biomarkers of liver injury or cancer in the plasma.

Associations of plasma proteins with haematological cancers (CLL, multiple myeloma, non-Hodgkin lymphoma) accounted for 20% (331/1,672) of all cancer type-specific associations despite these cancers comprising just 8% (463/6,073) of cancer cases evaluated, reflecting the origin of these malignancies in cells of the haematological system.

Among FDR-significant protein-cancer associations, we identified plasma proteins with established roles in cancer biology, clinical biomarkers for cancer early detection, and known therapeutic targets for cancer treatment, validating our approach. For example, the association with the strongest statistical evidence was for TNFRSF17 (also known as soluble BCMA), pharmacological target of the multiple myeloma monotherapy belantamab mafodotin, and multiple myeloma risk (HR 2.91 per SD increment, 95% CI 2.51-3.38, *q*-val = 4.102 x 10^−39^)(*34*). The second strongest association was for prostate-specific antigen (PSA), a widely used biomarker for prostate cancer detection, and prostate cancer risk (HR 1.98, 95% CI 1.78-2.20, *q*-val = 4.42 x 10^−32^). SEMA4A, a cell surface activator of T-cell mediated immunity and emerging cancer immunotherapy target, was strongly associated with risk of all three haematological cancers evaluated (CLL: HR 2.47, 95% CI 2.01-3.03, *q*-val = 1.45 x 10^−13^; multiple myeloma: HR 1.77, 95% CI 1.46-2.14, *q*-val = 7.34 x 10^−6^; non-Hodgkin lymphoma: HR 1.55, 95% CI 1.33-1.81, *q*-val = 1.48 x 10^−5^)(*35, 36*).

Along with validating known cancer biomarkers, we identified hundreds of previously unreported protein associations with cancer risk, including plasma proteins with strong and selective associations with single cancer types (GKN2 and stomach cancer: HR 1.50, 95% CI 1.31-1.70, *q*-val = 1.68 x 10^−6^; BPIFB1 and lung cancer: HR 1.42, 95% CI 1.24-1.62, *q*-val = 2.35 x 10^−4^), proteins exclusively expressed in the tissue of origin of the cancer evaluated (SERPINA7 and hepatocellular carcinoma: HR 2.48, 95% CI 1.79-3.41, *q*-val = 3.06 x 10^−5^; F9 and hepatocellular carcinoma: HR 0.44, 95% CI 0.32-0.62, *q*-val = 7.08 x 10^−4^), and targets of approved non-cancer medications (e.g. IL23, IL5RA, LAMC2). Together, these findings reveal a diverse catalogue of circulating proteins linked to cancer risk with potential relevance for disease biology, early detection, and drug repurposing.

### Confirmation analyses in ARIC

Of the 806 aptamers associated with risk of at least one cancer endpoint in EPIC that were taken forward to confirmation analyses, 581 had been measured on the SomaScan 5K Assay in ARIC, passed quality control procedures and were included for analysis. Overall, 54.8% of the 9,391 ARIC participants included in confirmation analyses were female and the median age at baseline was 57.5 years.

Among 697 aptamer–cancer associations tested for confirmation in ARIC, 78% (540/697) showed concordant directions of associations between EPIC and ARIC (**Fig. 4**). When considering concordance of findings at nominal significance in the ARIC study (one-tailed *P*<0.05), 48.6% (339/697) of aptamer-cancer associations identified in EPIC were replicated in the same direction in ARIC (**table S29**). Concordance between the studies differed markedly by cancer site (**fig. S4**). For example, more than 60% of the aptamers associated with CLL (57/93) and multiple myeloma (16/23) that were tested for replication in ARIC showed evidence of association in the same direction. In contrast, around 30% of the aptamers associated with lung (26/81) and post-menopausal breast cancer (3/10) that were tested for replication showed consistent evidence of association in ARIC.

**Fig. 4:**
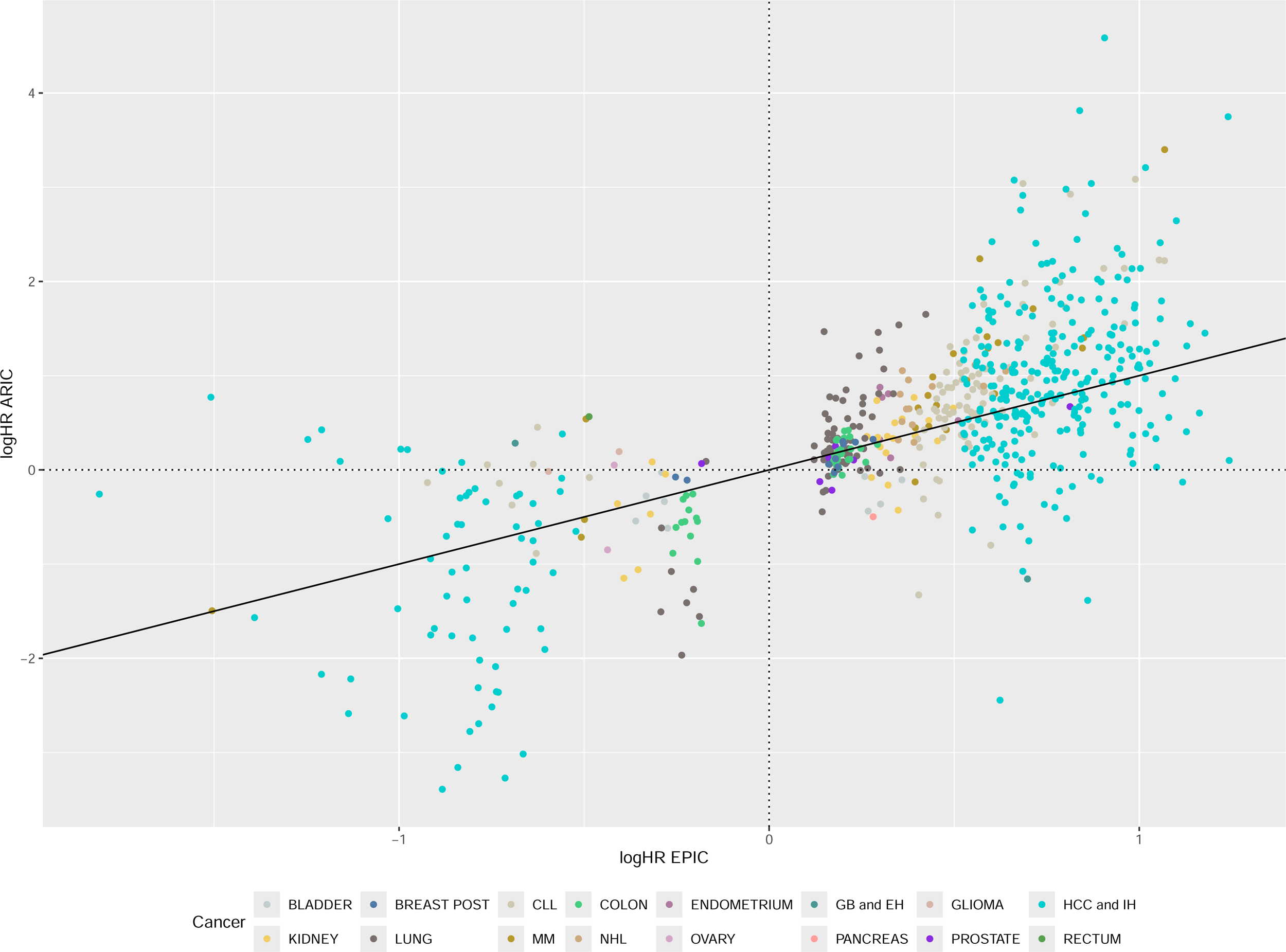
Comparison of logHRs of aptamer-cancer pairs across EPIC and ARIC.

### Analyses stratified by follow-up time

Examining FDR-significant protein-cancer associations by time since blood collection revealed that 76 protein-cancer associations were restricted to one of three follow-up periods evaluated (**tables S30-32**). This included 2 associations that were restricted to the first 2 years of follow-up including MMP12 and overall cancer (HR 1.36, 95% CI 1.20-1.53, *q*-val = 3.83 x 10^−4^) and 34 within the first 5 years of follow-up such as STC1 and kidney cancer (HR 2.63, 95% CI 1.99-3.49, *q*-val = 3.06 x 10^−8^), consistent with markers of sub-clinical disease and therefore of potential utility for cancer early detection. In addition, there were 40 associations identified in analyses excluding the first 5 years of follow-up such as OTC and hepatocellular carcinoma (HR 2.76, 95% CI 2.25-3.39, *q*-val = 3.31 x 10^−18^), indicating proteins more likely to reflect longer-term aetiological processes.

Notably, when expanding follow-up time stratified analyses to include protein-cancer associations that were not FDR-significant in primary analyses, we identified an additional 447 associations. Within the first 2 years after blood collection, we identified 19 additional protein–cancer associations including TFRC, a cell surface receptor that regulates cellular iron homeostasis and proliferation, which was strongly associated with colon cancer risk diagnosed within the first two years of follow-up (HR 1.95, 95% CI 1.47-2.59, *q*-val = 1.36 x 10^−3^) but not in analyses using complete follow-up time (HR 1.00, 95% CI 0.90-1.10, *q*-val = 0.99)(*37*). Likewise, GOLM1, a type 2 transmembrane protein found in the Golgi apparatus that is overexpressed in prostate cancer, was only associated with prostate cancer risk among cases diagnosed in the first two years of follow-up (HR 1.92, 95% CI 1.41-2.63, *q*-val = 8.46 x 10^−3^)(*38*).

Expanding analyses to the first 5 years of follow-up across all cancer types identified 315 additional protein–cancer associations not detected in analyses using complete follow-up, highlighting proteins with potential relevance as markers for near-term risk. For example, F9, a protein essential for blood clotting, was associated with kidney cancer risk in the first five years of follow-up (HR 3.45 95% CI 1.78-6.68, *q*-val = 2.16 x 10^−2^) but not in analyses using complete follow-up time (HR 1.24, 95% CI 0.99-1.55, *q*-val = 0.53)(*39*). Likewise, IFNB1, a cytokine that plays a critical role in regulating the immune response, was only associated with multiple myeloma risk in the first five years of follow-up (HR 0.12 95% CI 0.05-0.31, *q*-val = 2.81 x 10^−3^)(*40*). In total, when restricting analyses to the first five years of follow-up, 79 additional associations were identified for CLL, along with 65 for oesophageal squamous cell carcinoma and 57 for kidney cancer, representing 35% and 58% increases in the numbers of FDR-significant associations for these latter two cancers, respectively, as compared to those identified in analyses using complete follow-up time.

By contrast, excluding the first 5 years of follow-up identified 113 additional protein-cancer associations that were not apparent in analyses using complete follow-up. For example, TAGLN, an actin-binding protein that is down-regulated in prostate cancer tissue, was not associated with prostate cancer risk in analyses using complete follow-up time (HR 1.01, 95% CI 0.92-1.12, *q*-val = 0.96) but was strongly inversely associated with risk after excluding the first five years of follow-up (HR 0.57, 95% CI 0.48-0.68, *q*-val = 2.43 x 10^−7^)(*41*). Likewise, the cytokine and growth factor PTN was only associated with endometrial cancer risk (HR 0.47, 95% CI 0.37-0.60, *q*-val = 6.82 x 10^−7^) in analyses excluding the first five follow-up years(*42*). Around 50% of the 113 additional associations detected were for non-squamous cell carcinoma of the upper aerodigestive tract (20), overall cancer (12), hepatocellular carcinoma (12), and oesophageal squamous cell carcinoma (11). These findings highlight proteins with potential aetiological roles in cancer that emerge only after longer periods of follow up and merit further investigation to clarify their role in cancer development.

An overview of the number of protein associations across each follow-up time stratified analysis and each cancer type is presented in **Fig. 5**. Complete findings from follow-up time stratified analyses are presented in **table S33-35**.

**Fig. 5:**
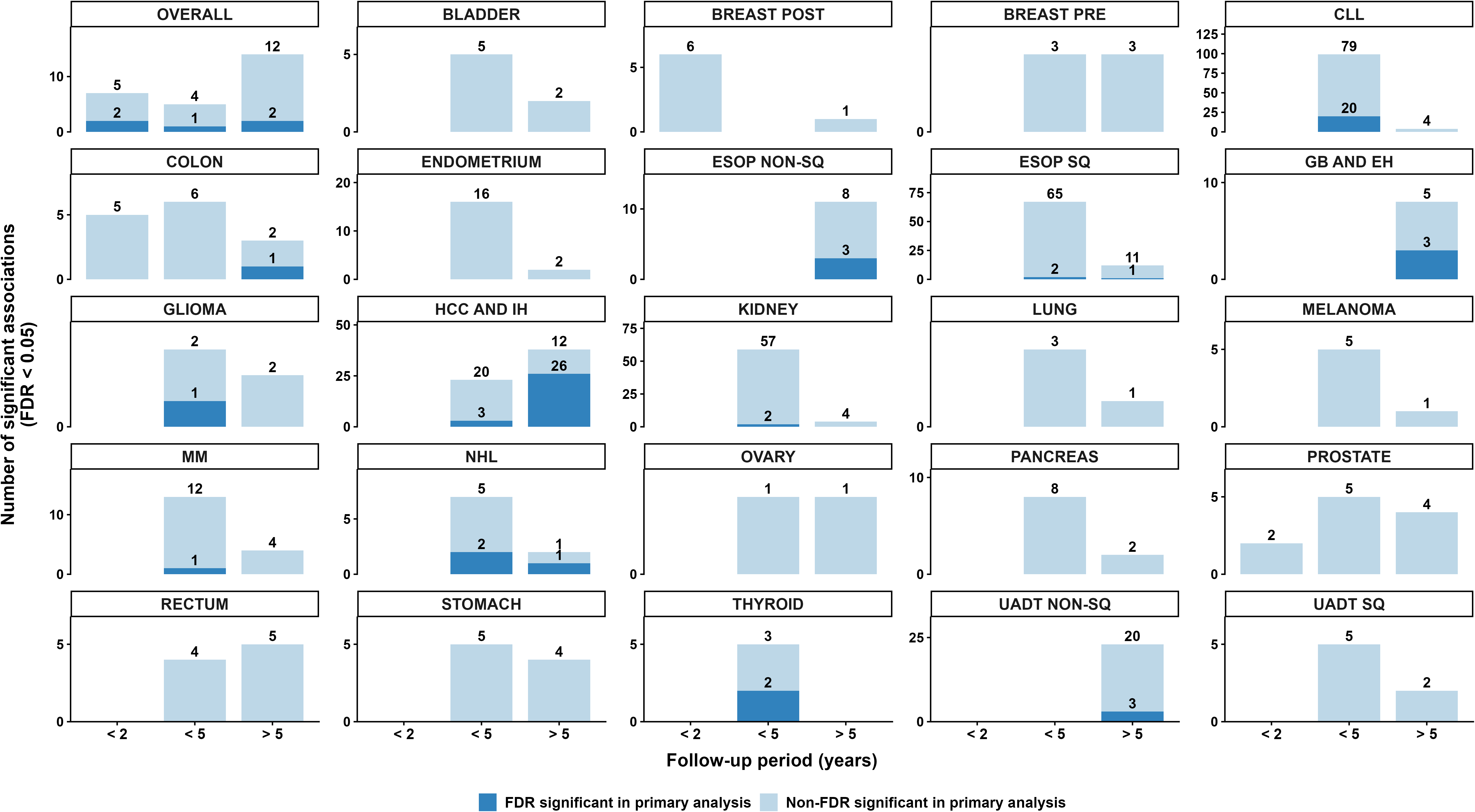
Overview of the number of FDR-significant protein associations across each follow-up time stratified analysis and each cancer type. Asterisk (*) denotes FDR-significance.

### Biological characterisation of prioritised proteins through over-representation analysis, tissue specificity analysis, and functional annotation

To gain potential mechanistic insights into the role of proteins linked to one or more cancer types, we employed over-representation enrichment analysis comparing FDR significant proteins from complete follow-up analyses against all SomaScan 7K proteins using three complementary resources (Reactome, Gene Ontology [GO], Human MsigDB Collections Hallmark gene set). In over-representation analysis using the Reactome resource in clusterProfiler, cancer-associated proteins were enriched for diverse biological pathways, including those related to regulation of the insulin-like growth factor (IGF) transport and uptake by insulin-like growth factor binding proteins (IGFBPs), diseases of metabolism, and other semaphorin interactions, along with 4 other pathways. Among the 22 enriched GO Biological processes (*q*-val < 0.05), we found enrichment for glycosaminoglycan binding, sulfur compound binding, and extracellular matrix binding. In over-representation analysis using the Human MSigDB Collections Hallmark gene set, proteins were enriched for gene sets implicated in xenobiotic metabolism, epithelial mesenchymal transition (EMT), interleukin-2/STAT5 signalling, and glycolysis. Genes encoding plasma proteins were highly expressed across a diverse range of tissues including subcutaneous and visceral adipose tissue, lung, breast, and small intestine (**fig. S5**). As compared to proteins not associated with risk of any cancer type, proteins associated with risk of one or more cancer types were more likely to be secretory proteins (37% [492/1,318] vs 21% [1,078/5,084], *P* = 1.19 x 10^−33^) and membrane proteins (29% [376/1,318] vs 23% [1,190/5,084], *P* = 1.34 x 10^−4^) but less likely to be intracellular proteins (71% [933/1,318] vs 79% [4,022/5,084], *P* = 1.55 x 10^−10^). Complete results from over-representation analyses are presented in **tables S36-38**.

### Association of proteins with overall cancer risk

Of the 60 proteins associated with overall cancer risk in multivariable-adjusted models, 56 were positively associated and four were inversely associated with cancer risk (STK4: HR 0.90, 95% CI 0.86-0.95, *q*-val = 0.03; NME4: HR 0.88, 95% CI 0.83-0.95, *q*-val = 0.04; CLEC3B: HR 0.92, 95% CI 0.87-0.96, *q*-val = 0.04; PGAM2: HR 0.91, 95% CI 0.87-0.96, *q*-val = 0.04). The strongest associations were observed for PSA (HR 1.20, 95% CI 1.14-1.27,*q*-val = 9.33 x 10^−9^), TFF3 (HR 1.15, 95% CI 1.10-1.20, *q*-val = 1.03 x 10^−5^), GPR37 (HR 1.14, 95% CI 1.08-1.19, *q*-val = 1.41 x 10^−4^), and MUC16 (HR 1.13, 95% CI 1.08-1.18, *q*-val = 1.69 x 10^−4^). TFF3 is a secretory protein that is widely expressed across several cancer types and has been shown to promote cell proliferation, invasion, and angiogenesis(*43, 44*). GPR37 regulates oligodendrocyte differentiation and central nervous system myelination and has been linked to Parkinson’s disease though the role of this protein in cancer risk is less established(*45*). MUC16 (also known as CA-125), a protein expressed in a variety of normal and cancer tissues, is an established diagnostic biomarker for ovarian cancer(*46, 47*). In addition to these proteins, we also observed several associations of members of the insulin-like growth factor (IGF) family (IGFBP4, IGFLR1) and inflammatory cytokines (IL23, IL18BP) with overall cancer risk. Fifteen proteins linked to overall cancer risk, including IL12B and TNFRSF1B, are targets of approved drugs, warranting further evaluation of their aetiological role in cancer development to inform potential drug repurposing opportunities for cancer prevention.

Around 55% (33/60) of proteins showed evidence of heterogeneity of association by cancer type (*P*_het_ < 0.05), suggesting that findings for overall cancer risk were driven by one or a small number of cancer types (**Fig. 6**). This included PSA where cancer type-specific associations were only detected for prostate and lung cancer, RARRES2 which was only associated with kidney and colon cancer, and CSNK2A1 which was exclusively associated with melanoma. Of the remaining 27 proteins, including TFF3, FCRL3, CLEC3B, GLYAT, and IGFLR1, the limited evidence of heterogeneity suggested potential shared roles of these proteins as causes, consequences, or (e.g. confounded) markers of cancer across multiple cancer sites. There were 9 proteins that associated with overall cancer risk at *q*-val <0.05 despite not being associated at FDR significance with any of the individual cancer types (PRC1, ILF3, PGAM2, ITLN1, TRIM5, TEX30, STK4, PI3, and PRSS2), suggesting proteins with possible shared but relatively weaker associations across cancer types and highlighting the benefit of examining proteins in relation to overall cancer risk.

**Fig. 6:**
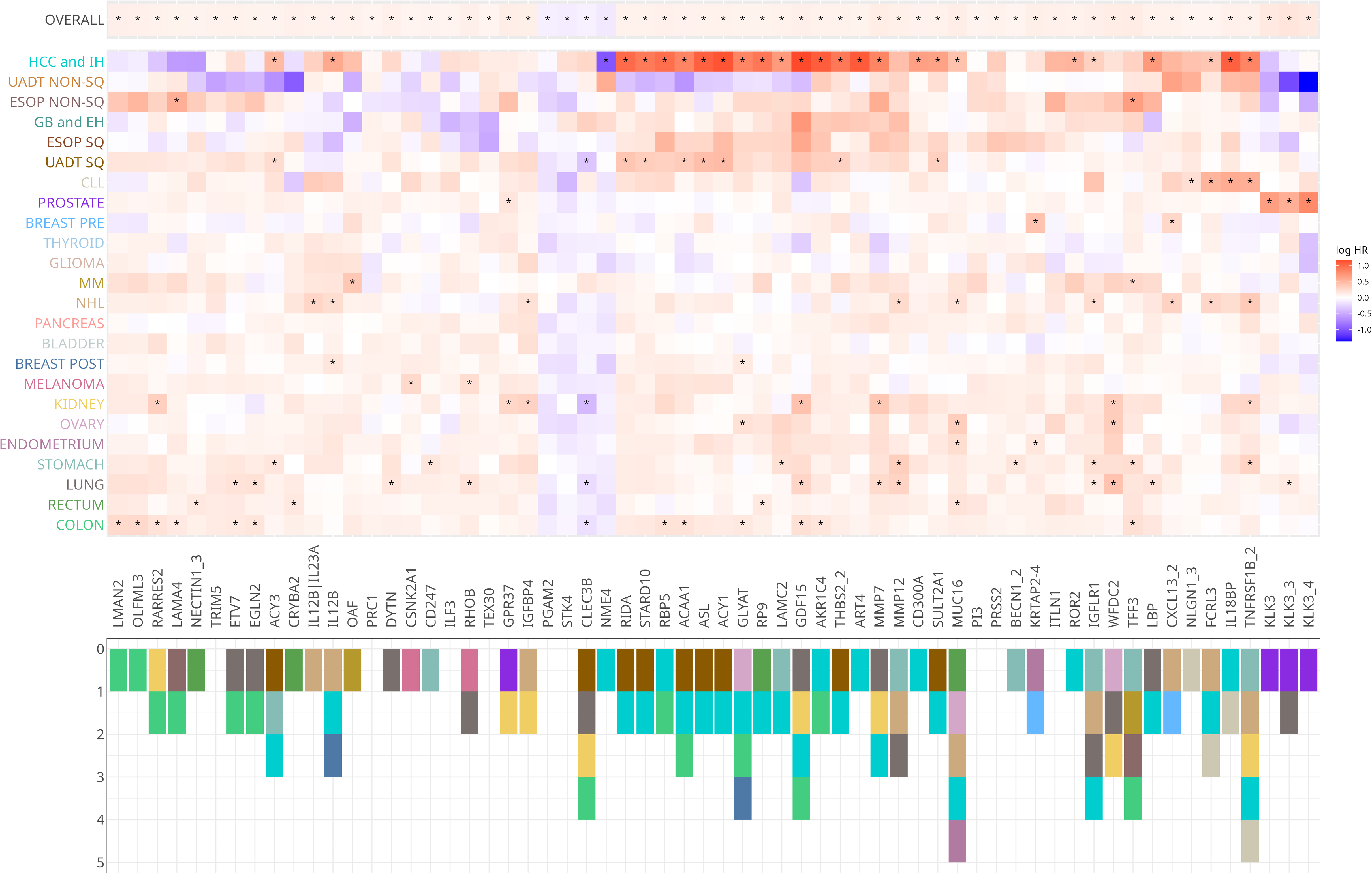
Overview of the strength and magnitude of association of each aptamer-cancer pair with FDR-significant evidence for association with overall cancer across individual cancer types.

### Sex-specific associations

Sex interaction analyses were conducted for non–sex-specific cancers only (i.e., excluding breast, prostate, ovarian and endometrial cancers). Across these analyses, there were 36 protein-cancer associations with evidence of interaction by sex (*q*-val < 0.05), 36% (13/36) of which were for glioma risk. For example, there was evidence of interaction of the association of SNX27 with glioma risk (*q*-val = 8.78 x 10^−3^) with evidence of a positive risk relationship in men (HR 1.40, 95% CI 1.07-1.83) and a protective association in women (HR 0.44, 95% CI 0.31-0.62). ALPL, which contributes to skeletal mineralisation, also showed evidence of a sex-specific association with overall cancer risk (HR men: 1.14, 95% CI 1.05-1.23, HR women: 0.89, 95% CI 0.83-0.95, *q*-val = 3.62 x 10^−2^).

### Proteins associated with multiple cancer sites

A total of 284 proteins were associated with risk of two or more cancer types, with 13 associated with 4 or more including MUC16, TNFRSF1B, PIGR, CD48, and GDF15. MUC16/CA-125, an antigenic tumour marker expressed by ovarian cancer cells along with cells lining various organs including the endometrium, fallopian tubes, colon, kidney, and stomach, is an established diagnostic marker for ovarian cancer. In addition to ovarian cancer risk, we identified positive associations of MUC16/CA-125 with risk of hepatocellular carcinoma, endometrial cancer, non-Hodgkin lymphoma, and rectal cancer, consistent with prior studies that have identified potential roles for this protein as a biomarker for early detection and risk stratification for hepatocellular carcinoma, endometrial cancer, and other non-ovarian cancers(*48–50*). TNFRSF1B, a member of the TNF receptor superfamily that mediates the effect of TNF in inflammation and autoimmunity, was associated with all three haematological cancers evaluated along with cancers of the liver, stomach, and kidney(*51*). Both CLSTN1, a transmembrane protein that has been implicated in cell proliferation and CD48, a member of the signalling lymphocyte activation molecular family, were positively associated with risk of all three haematological cancers and hepatocellular carcinoma(*52, 53*). GDF15, a marker of cellular stress with widespread pleiotropic associations across several cardiometabolic, autoimmune, and neurological conditions was associated with liver, kidney, lung, and colon cancer risk(*15, 54–57*). LAG3, the target of the immunotherapy relatlimab used to treat advanced melanoma, was positively associated with risk of CLL, non-Hodgkin lymphoma, and hepatocellular carcinoma, highlighting the potential for repurposing of this medication to novel indications(*58*). Cancer types that shared the most proteins were hepatocellular carcinoma and squamous cell carcinoma of the upper aerodigestive tract (64), CLL (31), and non-Hodgkin lymphoma (31); CLL and non-Hodgkin lymphoma (24); and hepatocellular carcinoma and oesophageal squamous cell carcinoma (17).

### Mapping of cancer-linked proteins to approved medications

Overall, 315 proteins (measured by 347 aptamers) associated with risk of at least one cancer type were also targets of one or more approved drugs in DrugBank or ChEMBL (**table S39**). Over 50% of all associations between proteins targeted by medications and cancer risk were for hepatocellular carcinoma (37%, 174/474), CLL (9%, 43/474), or squamous cell carcinoma of the upper aerodigestive tract (8%, 37/474). Notably, the protein targets MUC16/CA-125, TNFRSF1B, GLYAT, and CLEC3B were associated with risk of 5 or more cancer types (including overall cancer).

Proteins mapped to drugs included targets of approved cancer medications such as PARP1 (i.e. target of niraparib, associated with risk of CLL) and EGFR (target of gefitinib, associated with risk of stomach and squamous cell carcinoma of the oesophagus) along with targets of non-cancer medications such as IL12B (target of ustekinumab, used to treat autoimmune conditions, associated with risk of non-Hodgkin lymphoma, hepatocellular carcinoma, and post-menopausal breast cancer) and CALCB (target of galcanezumab, used in migraine disorder, associated with risk of multiple myeloma), highlighting a large array of potential drug repurposing opportunities that require further verification in independent studies(*59–62*). We also found instances where proteins associated with cancer risk at specific cancer sites were mapped to drugs that are approved to treat these cancers including SLAMF7 (target of elotuzumab, used to treat multiple myeloma) and EPHA2 (target of regorafenib, used to treat hepatocellular carcinoma), suggesting shared roles of these proteins across different stages of the carcinogenesis spectrum for these cancers.

## Discussion

Our large-scale profiling of circulating proteins within a prospective cohort revealed a vast landscape of previously unrecognised plasma biomarkers linked to cancer development. Across 25 cancer outcomes, we identified 1,732 protein-cancer associations, many of which have not previously been implicated in cancer risk. While our analyses recapitulate a set of established biomarkers, we also significantly expand the number of plasma proteins associated with incident cancer, illuminating novel biological pathways involved in carcinogenesis and highlighting candidate targets for risk stratification, early detection, and therapeutic intervention.

The scale of this investigation, encompassing proteomic measurements of more than 6,400 unique circulating proteins in over 10,000 participants, including 6,073 incident cancer cases across 25 cancer outcomes, enabled the identification of approximately 50% more protein– cancer associations than the largest previous prospective pan-cancer proteomic analysis (i.e. 1,099 associations from an analysis of 2,920 plasma proteins with risk of cancer in 4,873 incident malignant cases at *P* < 1 x 10^−5^), including substantial expansions in the number of proteins associated with oesophageal (110 vs 12), ovarian (39 vs 1), bladder (39 vs 1), and stomach cancers (32 vs 5)(*55*). We replicate several previously reported associations, such as HAVCR1 with kidney cancer and CXCL13 with non-Hodgkin lymphoma, but not others, such as PLAUR with lung cancer, which could reflect differences in study populations, risk factors, confounder adjustment strategies, and/or proteomics platforms employed(*55*).

Stratifying analyses by time from blood collection revealed pronounced temporal structure in protein-cancer associations, highlighting 76 associations from our primary analyses that were driven by signals within one of three follow-up periods evaluated. When exploring protein-cancer pairs that were not FDR-significant in primary analyses, an additional 447 associations emerged only within specific follow-up windows, suggesting distinct proteomic signals across the carcinogenic trajectory, from markers of sub-clinical disease (<2 years) to near-term risk (<5 years) and proteins more consistent with longer-term aetiological processes (>5 years). This temporal stratification suggests that distinct biological pathways are activated across different stages of carcinogenesis, highlighting potential windows during which specific molecular targets or circulating biomarkers may become informative for early detection or therapeutic intervention.

Functional enrichment analyses indicated that proteins linked to cancer risk participate in diverse biological processes, including epithelial–mesenchymal transition, insulin-like growth factor-related pathways, glycosaminoglycan binding, complement cascades, and xenobiotic metabolism. Proteins associated with at least one cancer type were more likely to be members of the secretome or membrane-bound proteins, suggesting an enrichment for cancer risk associations for bioactive molecules that are actively secreted to peripheral blood to exert physiological functions and for proteins that may enter the circulation via cell death or injury(*63*). Notably, 315 of the identified proteins are targets of approved drugs, highlighting potential opportunities for therapeutic repurposing and preventive interventions, in particular where the causal relevance of these proteins can be further established using genetic epidemiological approaches such as *cis*-Mendelian randomization(*64*). Where these proteins are actively released into the circulation from non-cancerous tissue and exert downstream functional effects on tissues that give rise to cancer, they are more likely to reflect protein targets amenable to cancer prevention, which can be further interrogated in future studies.

Most proteins identified in this study were associated with only a single cancer type at FDR-significance, suggesting largely distinct circulating proteomic profiles across tumour types. In addition, of the 284 proteins associated with multiple cancers, 220 (77%) were associated with two cancer types. These findings highlight potential challenges when attempting to design therapeutic interventions or early detection tools aimed at preventing or detecting multiple cancer types. Nonetheless, the select proteins that were associated with several cancer types, including those with roles in cancer that are less characterised such as SVEP1 and GLYAT, can provide a unique window into possible shared biological mechanisms across multiple cancer types that merit further investigation. Several of these proteins have established roles in inflammation and immune signalling (e.g. TNFRSF1B, CD48, CD14, IL12B) and EMT (e.g. PLAUR, VCAM1, QSOX1). These shared associations suggest that systemic inflammatory and immune pathways, as well as processes related to cellular plasticity and tissue remodelling, may contribute to tumour development across multiple tissues. Eight proteins including PRC1, ILF3, and PGAM2 were strongly associated with overall cancer risk despite not showing FDR-significant associations with individual cancer types, suggesting shared but modest effects across tumour types that may be relevant for pan-cancer risk prediction or prevention strategies.

Strengths of this analysis include the comprehensive prospective assessment of > 6,400 proteins in relation to overall and site-specific cancer risk in a large case-cohort, the majority of which have not been previously evaluated across a large number of cancer sites in a prospective setting. The extensive phenotyping available within EPIC permitted comprehensive cancer type-specific adjustment for potential confounders. The long follow-up permitted evaluation of differential associations of proteins with cancer risk by follow-up time, highlighting proteins that were associated with cancer risk in specific follow-up time periods only that may have with potential utility as early detection, or aetiological biomarkers of cancer development. In addition, we were able to explore the potential transportability of our findings in an independent prospective cohort where we showed general directional concordance of hazard ratios across studies. Though both studies employed similar confounder adjustment strategies, they differed in participant characteristics, plasma collection tube types, and median follow-up time which could account for some of the differences observed for select aptamer-cancer comparisons.

Our study has several limitations. First, though these analyses represent the largest prospective investigation of plasma proteins and cancer risk to date, the number of cancer cases was limited for certain rarer cancer types. Second, we identified several instances where only one of multiple aptamers that bind to the same protein were associated with cancer risk which could reflect the binding of aptamers to specific protein conformations or isoforms or differences in their cross-reactivity, complicating interpretation of findings(*65*). Third, we did not stratify cancer type-specific analyses on histological grade or stage which may show differing associations with proteins evaluated. Fourth, though analyses were adjusted for a comprehensive set of cancer type-specific confounders, we cannot rule out the presence of confounding due to unmeasured factors (e.g. metabolic dysfunction-associated steatotic liver disease for hepatocellular carcinoma) or residual confounding (i.e. due to measurement error), which can limit conclusions into the aetiological role of proteins identified in this analysis. Finally, our primary analyses were restricted to individuals of European ancestry. Though 24% of participants in confirmation analyses in ARIC were of African ancestry, our findings require independent replication in other large cohorts of participants with non-European ancestry to further inform on transportability of findings across ancestry groups.

Our findings provide a comprehensive map of circulating proteins associated with cancer risk across multiple tumour types and stages of disease development. The identification of hundreds of previously unrecognised protein–cancer associations, including many corresponding to existing drug targets, highlights opportunities to advance blood-based strategies for cancer early detection, risk stratification, prevention, and drug repurposing. By integrating large-scale proteomic profiling with a large population-based study, we were able to identify not only cancer-specific and shared protein markers and associated pathways, but also to characterise how these signals evolve across the natural history of disease. Such insights are critical for identifying when key biological pathways become dysregulated during carcinogenesis and for prioritising biomarkers and therapeutic targets most relevant to different stages of cancer development. Together, these results position the circulating proteome as a powerful resource for understanding cancer development and for informing next-generation approaches to cancer prevention and early detection.

## Supporting information

Supplementary Results

## Acknowledgments

We thank all EPIC participants for donating the samples that enabled our research. The authors thank the National Institute for Public Health and the Environment (RIVM), Bilthoven, the Netherlands, for their contribution and ongoing support to the EPIC Study. The authors are grateful to Clare Paterson and Hannah Biegel (Standard BioTools) for helpful feedback on an earlier version of this manuscript. The authors are also grateful to Rawan M Maawadh and Isobel G Jackson for their support in surveilling the plasma protein-cancer epidemiological literature for an earlier version of this manuscript. Cancer incidence data have been provided by the Maryland Cancer Registry, Center for Cancer Surveillance and Control, Department of Mental Health, and Hygiene, 201 W. Preston Street, Room 400, Baltimore, MD 21201. We acknowledge the State of Maryland, the Maryland Cigarette Restitution Fund, and the National Program of Cancer Registries (NPCR) of the Centers for Disease Control and Prevention (CDC) for the funds that helped support the availability of the cancer registry data.

## Author contributions

Conceptualization and study design: MJG, ER; Data analysis: VV, VAB, JY; Collected, prepared, analyzed, and/or contributed samples to the study: VV, VAB, DCM, PMK, CML, KT, PF, MDCL, AJ-Z, RT, CS, NW, ZH, KS-B, RT, NC, EAP, RCHV. Writing – original draft: JY, VV, MJG; Writing – review & editing: All authors. All authors read and approved the final manuscript.

## Data, code, and materials availability

EPIC data are available for investigators who seek to answer important questions on health and disease in the context of research projects that are consistent with the legal and ethical standard practices of IARC/WHO and the EPIC Centres. For information on how to submit an application for gaining access to EPIC data and/or biospecimens, please follow the instructions http://epic.iarc.fr/access/index.php.

Pre-existing data access policies for the parent ARIC cohort study specify that research data requests can be submitted to the steering committee; these will be promptly reviewed for confidentiality or intellectual property restrictions and will not be refused unreasonably. Individual level patient or protein data may further be restricted by consent, confidentiality, or privacy laws/considerations. These policies apply to both clinical and proteomic data. Additional information on how to obtain data, including access to some data via BioLINCC, which is free of charge and without the need for the ARIC Study approval, is available at: https://aric.cscc.unc.edu/aric9/researchers/Obtain_Submit_Data.

## Ethics approval and consent to participate

The EPIC study was conducted in accordance with the Declaration of Helsinki. The study was approved by the local ethical committees in participating countries and the IARC ethical committee. All participants provided written informed consent for data collection and storage, as well as individual follow-up before study entry. Approval for the ARIC study was received from the Institutional Review Board at each study center. All participants provided informed consent.

## IARC Disclaimer

Where authors are identified as personnel of the International Agency for Research on Cancer/World Health Organization, the authors alone are responsible for the views expressed in this article and they do not necessarily represent the decisions, policy, or views of the International Agency for Research on Cancer/World Health Organization.

## Supplementary Materials

Supplementary Methods

Supplementary Results

Figs 1-6

Supplementary Figs 1-5

Supplementary Tables 1-39

---

**fig. S1:**
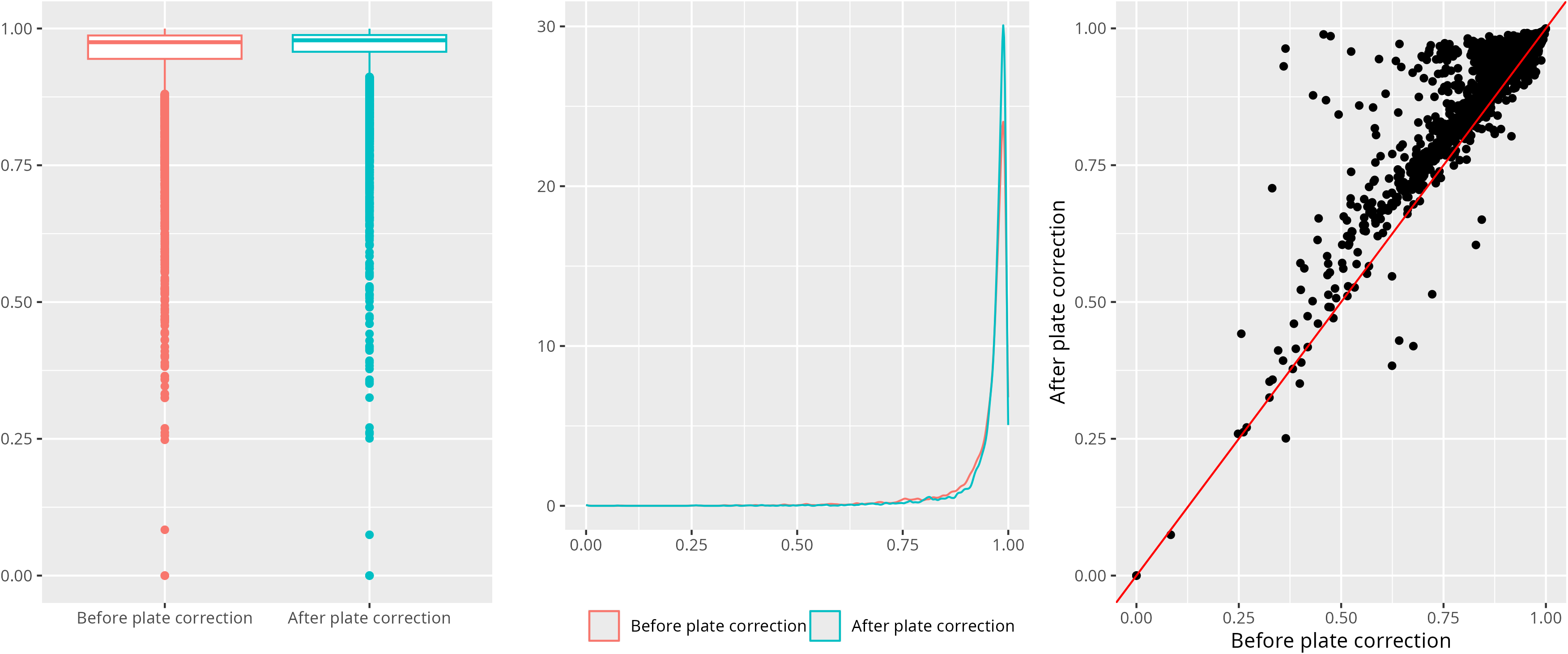
Analyses examining the impact of plate correction on assay reproducibility.

**fig. S2:**
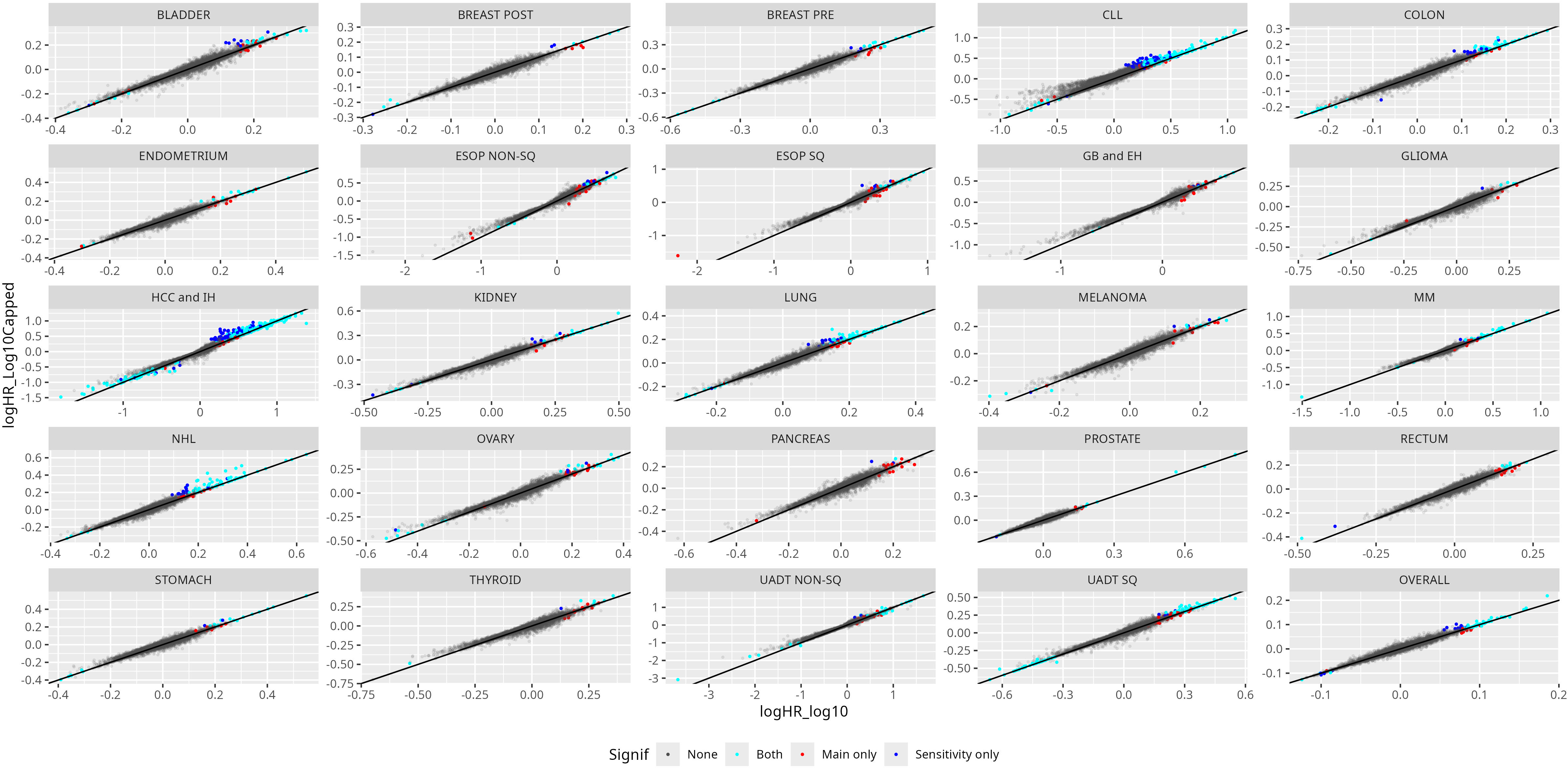
Analyses examining the impact of capping log-transformed relative abundances at greater or less than 5 standard deviations from the mean.

**fig. S3:**
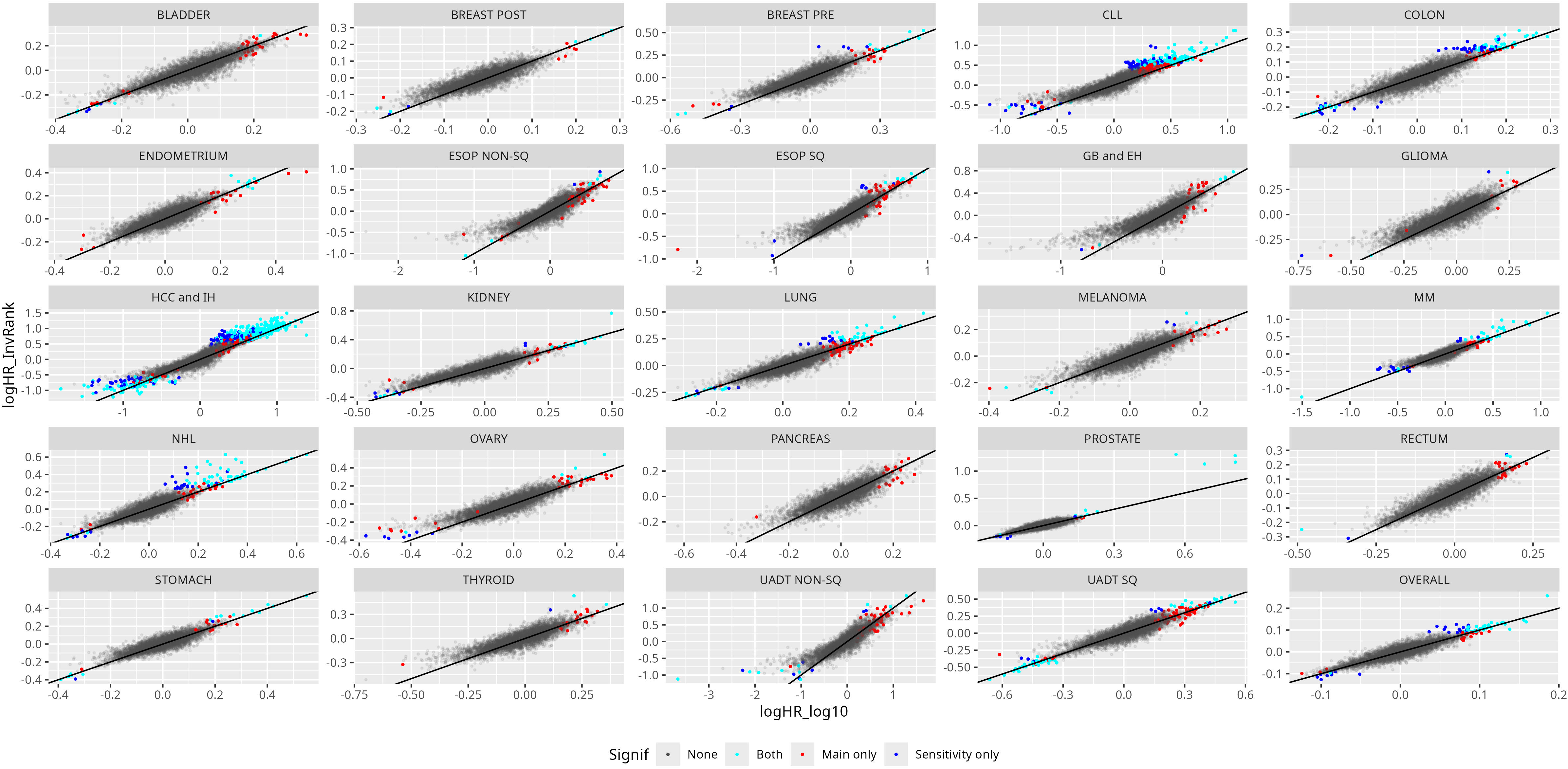
Analyses examining the impact of using inverse-rank normalization instead of log-transformation.

**fig. S4:**
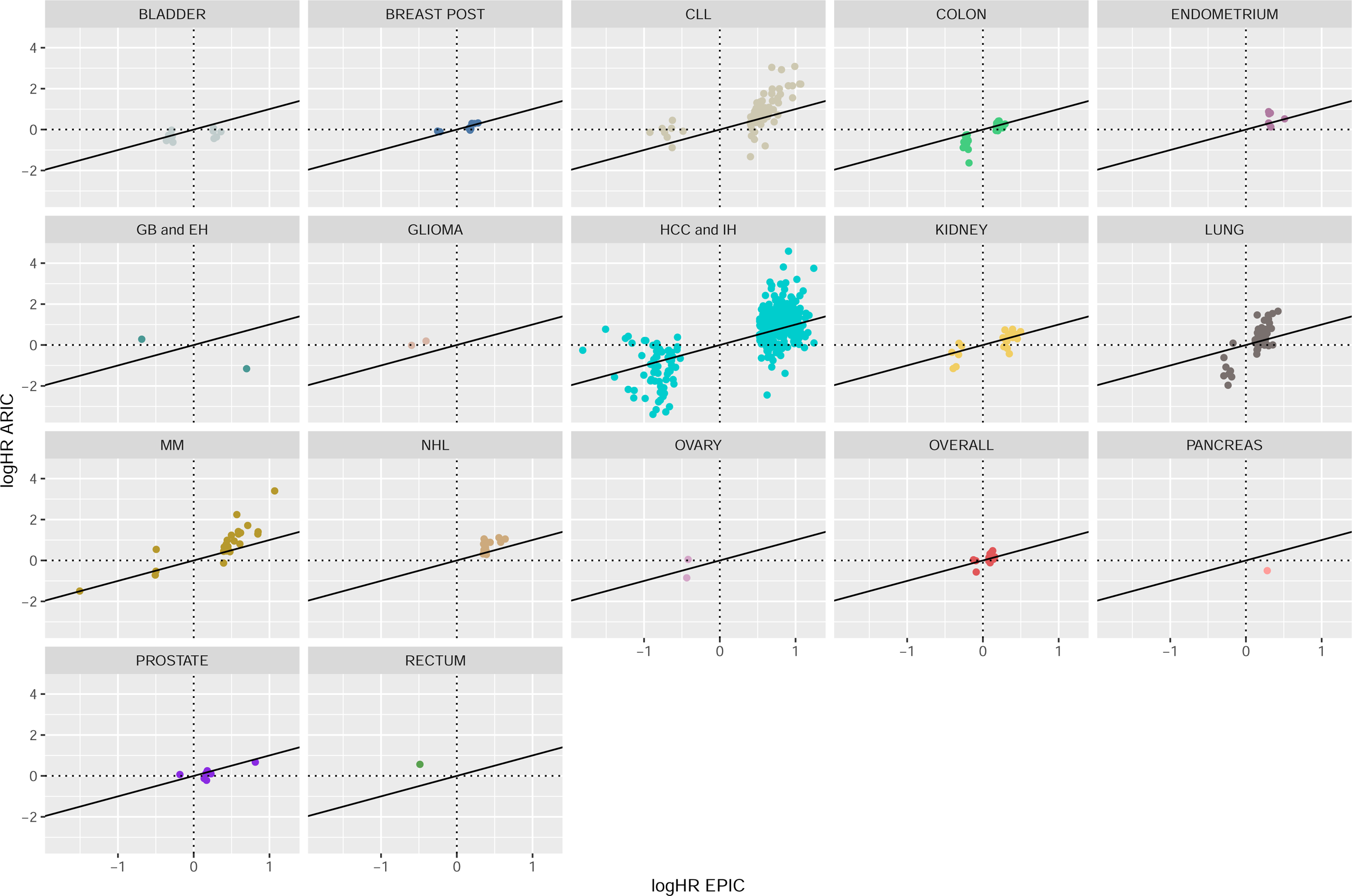
Comparison of logHRs of aptamer-cancer pairs across EPIC and ARIC per cancer type.

**Fig. S5:**
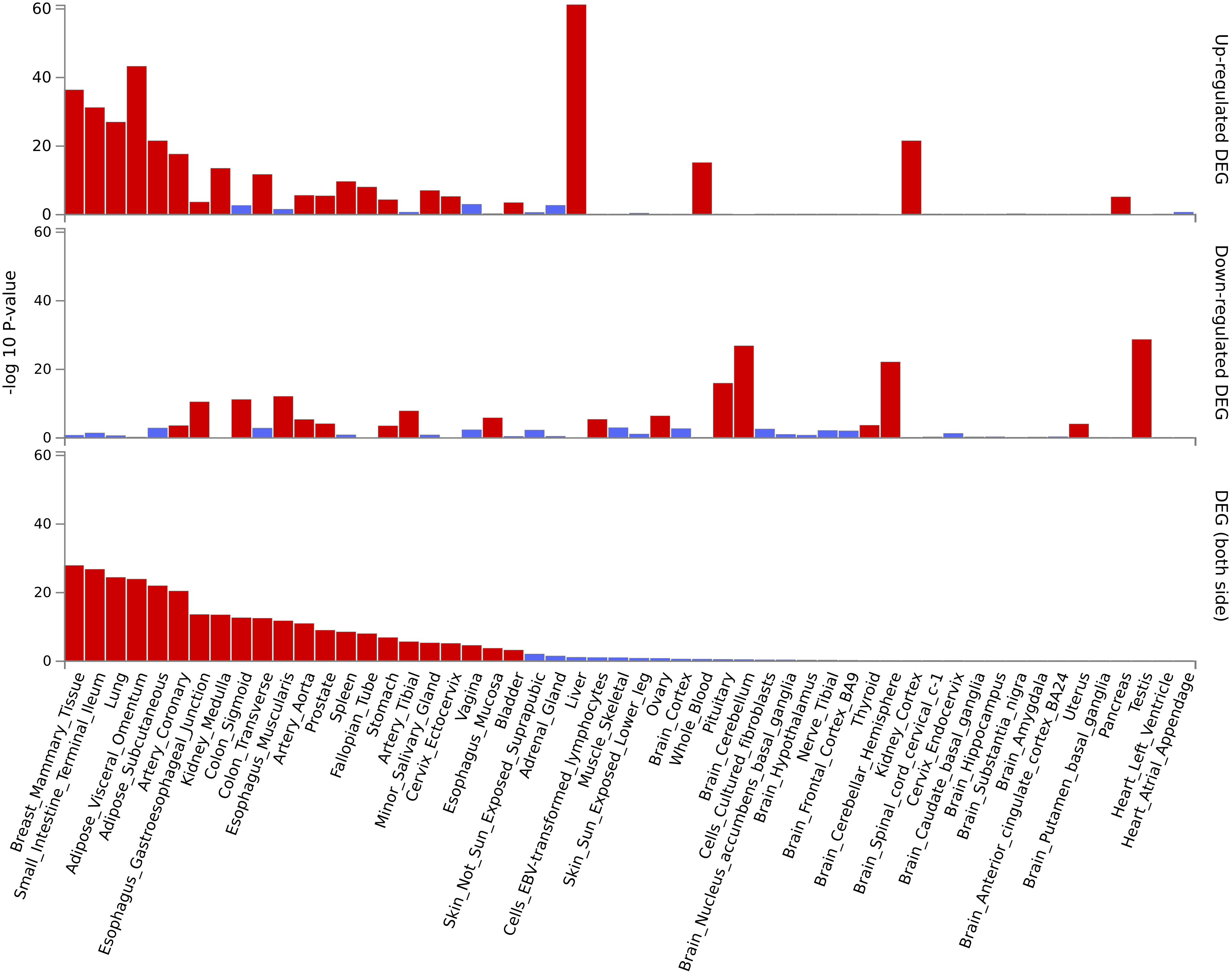
Tissue-specific differentially expressed gene analysis as performed in FUMA. Significantly enriched DEG sets (Pbon < 0.05) are highlighted in red.

