## Supplementary Results for "Population-scale plasma proteomics reveals pre-diagnostic biomarkers and shared pathways of cancer risk across 25 cancers"

Cancer type-specific analyses

*Liver hepatocellular carcinoma*

A total of 502 proteins (548 aptamers) were associated with risk of hepatocellular carcinoma (HCC), the most common form of primary liver cancer, including ALDH1A1 (HR 2.64, 95% CI 2.10-3.30, *q*-val = 6.98 x 10^-13^), CNDP1 (HR 0.47, 95% CI 0.39-0.56, *q*-val = 9.35 x 10^-13^), and NFASC (HR 2.06, 95% CI 1.73-2.44, *q*-val = 1.58 x 10^-12^). Around 20% (98/502) of proteins linked to HCC had “enriched” or “enhanced” expression within the liver. Cognate genes for HCC-associated proteins were highly upregulated in the liver and were enriched for Hallmark gene sets implicated in the EMT, glycolysis, and coagulation, among others. Among top protein associations, we identified associations with targets of approved and emerging HCC medications (e.g. MET, BRAF) along with previously proposed diagnostic biomarkers for HCC (e.g. CD163)(*1-3*). We also identified novel plasma proteins associated with HCC risk including ASL (HR 2.89, 95% CI 2.24-3.75, *q*-val = 7.22 x 10^-12^), LGALS3BP (HR 2.88, 95% CI 2.22-3.73, *q*-val = 1.88 x 10^-11^), and OTC (HR 2.63, 95% CI 2.07-3.34, *q*-val = 2.11 x 10^-11^). An additional 12 associations (*q*-val < 0.05) were identified after excluding the first 5 years of follow-up, including ARFIP2 (HR 0.22, 95% CI 0.14-0.35, *q*-val = 1.20 x 10^-7^) and VWC2 (HR 0.40, 95% CI 0.30-0.54, *q*-val = 2.80 x 10^-6^), highlighting proteins with potential aetiological roles in cancer development that may only emerge after longer follow-up periods. 30 plasma proteins were associated with risk of gallbladder and extra-hepatic cancer including ETV5 (HR 1.42, 95% CI 1.27-1.58, *q*-val = 7.11 x 10^-7^), NKD2 (HR 1.30, 95% CI 1.17-1.44, *q*-val = 4.11 x 10^-4^), and C7orf69 (HR 1.21, 95% CI 1.12-1.30, *q*-val = 6.78 x 10^-4^).

*Haematological cancers*

In total, we identified 331 protein associations with risk of CLL, non-Hodgkin lymphoma (NHL), or multiple myeloma which largely overlap with protein associations identified in a recent analysis of lymphoid cancer subtypes in EPIC(*4*). Of these, 204 proteins were associated with CLL, the most common form of adult leukaemia, with the strongest associations identified for FCMR (HR 2.87, 95% CI 2.35-3.51, *q*-val = 3.46 x 10^-20^), an immunoglobulin receptor that is highly expressed on the surface of CLL B cells(*5*). We also observed strong associations for SEMA4A (HR 2.47, 95% CI 2.01-3.03, *q*-val = 1.45 x 10^-13^), and SELL (HR 2.69, 95% CI 2.14-3.39, *q*-val = 7.13 x 10^-13^) along with multiple cell surface markers (CD14, CD46, CD48, CD72) and members of the tumour necrosis factor receptor superfamily (TNFRSF1B, TNFRSF9, TNFRSF13B, TNFRSF14). 75 proteins were associated with NHL risk, including FCER2 (HR 1.90, 95% CI 1.66-2.17, *q*-val = 4.50 x 10^-16^), FCMR (HR 1.74, 95% CI 1.55-1.96, *q*-val = 4.50 x 10^-16^), and FCRL1 (HR 1.79, 95% CI 1.53-2.08, *q*-val = 4.90 x 10^-10^), all of which are highly expressed in B-cells from which the majority of NHL cases arise. We identified 52 proteins associated with multiple myeloma risk including TNFRSF17 (HR 2.91, 95% CI 2.51-3.38, *q*-val = 4.10 x 10^-39^), IL5RA (HR 1.80, 95% CI 1.58-2.06, *q*-val = 3.57 x 10^-14^), and SLAMF7 (HR 2.33, 95% CI 1.89-2.88, *q*-val = 3.14 x 10^-11^). We also identified an additional 12 protein associations when analyses were restricted to the first five years of follow-up including SPAG7 (HR 1.32, 95% CI 1.21-1.44, *q*-val = 1.86 x 10^-7^) and HEBP1 (HR 0.65, 95% CI 0.54-0.78, *q*-val = 1.27 x 10^-3^). Notably, 31 proteins were shared across at least two of three haematological cancers with CD48, CLSTN1, LY9, and SEMA4A showing strong and positive associations across all three.

*Breast cancer*

13 proteins were associated with post-menopausal breast cancer risk, with the strongest associations observed for LEG1 (HR 1.32, 95% CI 1.18-1.49, *q*-val = 9.43 x 10^-4^), SAR1B (HR 1.22, 95% CI 1.12-1.32, *q*-val = 1.07 x 10^-3^), and CST6 (HR 1.30, 95% CI 1.15-1.46, *q*-val = 4.63 x 10^-3^). LEG1 is an evolutionary conserved protein potentially implicated in liver development that has an unclear role in breast cancer risk(*6*). Plasma protein LEG1 homolog levels have recently been reported to associate with an increased risk of post-menopausal breast cancer (HR 1.45, 95% CI 1.23-1.70 per doubling) in the ARIC study(*7*). Common variants in *CST6* have previously been linked to breast cancer risk in genome-wide association studies (GWAS), further validating CST6 as a potential aetiological biomarker in breast cancer development(*8, 9*). We also identified 29 proteins associated with pre-menopausal breast cancer risk, including KLK5 (HR 1.26, 95% CI 1.17-1.36, *q*-val = 2.85 x 10^-6^), ADPGK (HR 1.63, 95% CI 1.36-1.94, *q*-val = 5.29 x 10^-5^), and LACRT (HR 1.52, 95% CI 1.30-1.79, *q*-val = 1.62 x 10^-4^). Three proteins (LEG1, LACRT, PIP) were positively associated with risk of both post- and pre-menopausal breast cancer.

*Lung cancer*

116 plasma proteins were associated with lung cancer risk. Among top protein associations, we replicated previously reported prospective associations of select proteins (WFDC2, PIGR, CAPG) but also identify novel plasma proteins implicated in lung cancer risk including SCFD1 (HR 1.22, 95% CI 1.13-1.32, *q*-val = 1.26 x 10^-4^), POLE3 (HR 1.19, 95% CI 1.11-1.27, *q*-val = 2.38 x 10^-4^), and EGLN2 (HR 1.24, 95% CI 1.14-1.36, *q*-val = 7.98 x 10^-4^). Proteins associated with lung cancer risk were strongly enriched for GO biological processes related to the immune response including gene sets related to positive regulation of defense response and the innate immune response, among other pathways. We also identified an additional 3 plasma proteins associated with lung cancer risk when analyses were restricted to the first five years of follow-up (PKD2: HR 1.30, 95% CI 1.15-1.47, *q*-val = 4.06 x 10^-3^; AZGP1: HR 0.72, 95% CI 0.48-0.87, *q*-val = 1.11 x 10^-2^; ENAH: HR 1.83, 95% CI 1.31-2.56, *q*-val = 2.87x 10^-2^), highlighting proteins with potential utility as biomarkers for risk stratification or early detection of cancer that merit further investigation.

*Gastrointestinal cancers*

78 proteins were associated with colon cancer risk including previously reported protein biomarkers of colon cancer (TFF3), proteins encoded by known colon and/or colorectal cancer susceptibility genes (*INHBC*, *IQCF1*, *EHMT2*), and targets of emerging cancer immunotherapies (IFNG)(*10-13*). We also identified novel associations of plasma proteins with colon cancer risk including TCEAL8 (HR 1.22, 95% CI 1.13-1.31, *q*-val = 1.16 x 10^-4^), COMP (HR 0.78, 95% CI 0.71-0.86, *q*-val = 2.42 x 10^-4^), and OLFML3 (HR 1.31, 95% CI 1.18-1.45, *q*-val = 3.41 x 10^-4^) which has previously been shown to promote tumour angiogenesis and growth *in vivo(14)*. 20 proteins were association with rectal cancer risk, including NUDT1 (HR 0.61, 95% CI 0.51-0.75, *q*-val = 5.56 x 10^-4^), CSTB (HR 1.21, 95% CI 1.11-1.32, *q*-val = 2.27 x 10^-3^), and FRS2 (HR 1.16, 95% CI 1.09-1.25, *q*-val = 4.96 x 10^-3^). No proteins were shared across colon and rectal cancer risk.

We identified 48 proteins associated with oesophageal squamous cell carcinoma risk and 59 with oesophageal non-squamous cell carcinoma risk. FRS2, an adaptor protein that plays an important role in FGFR signalling, was the protein most strongly associated with oesophageal squamous cell carcinoma risk (HR 1.23, 95% CI 1.15-1.31, *q*-val = 3.81 x 10^-6^). When analyses were restricted to the first five years of follow-up, an additional 65 proteins not identified in primary analyses were associated with oesophageal squamous cell carcinoma including ANXA3 (HR 1.63, 95% CI 1.42-1.86, *q*-val = 2.26 x 10^-9^), HNRNPA2B1 (HR 0.12, 95% CI 0.05-0.26, *q*-val = 7.89 x 10^-5^), and TCP11L1 (HR 1.38, 95% CI 1.22-1.56, *q*-val = 9.73 x 10^-5^). The strongest association for risk of non-squamous cell carcinoma of the oesophagus was for BCL6 (HR 1.50, 95% CI 1.32-1.71, *q*-val = 1.01 x 10^-6^), a transcriptional repressor required for the differentiation of T follicular helper (Tfh) cells that has previously been implicated in B cell lymphomas and breast cancer(*15, 16*). We also found a strong association of ANGPT4, an adipokine shown to promote invasion and metastasis across several cancer sites, with risk of non-squamous cell carcinoma of the oesophagus (HR 1.42, 95% CI 1.24-1.63, *q*-val = 1.80 x 10^-4^)(*17*).

32 proteins were associated with stomach cancer risk, most notably REG4 (HR 1.73, 95% CI 1.47-2.05, *q*-val = 2.31 x 10^-7^), which has been shown to increase the size of gastric tumours in mice and to promote the proliferation of gastric cancer cells, and GKN2 (HR 1.50, 95% CI 1.31-1.70, *q*-val = 1.68 x 10^-6^), an anti-inflammatory protein that is highly expressed in the gastric epithelium(*18-20*). We also identified novel protein associations with stomach cancer risk including DDAH1 (HR 1.33, 95% CI 1.18-1.50, *q*-val = 1.36 x 10^-3^), TCEAL4 (HR = 1.21, 95% CI 1.12-1.31, *q*-val = 1.49 x 10^-3^), and EGFR (HR 0.68, 95% CI 0.57-0.80, *q*-val = 1.73 x 10^-3^).

17 proteins were associated with pancreatic cancer risk with the strongest associations observed for RIPPLY1 (HR 1.20, 95% CI 1.11-1.29, *q*-val = 9.19 x 10^-4^), FSTL5 (HR 1.23, 95% CI 1.12-1.35, *q*-val = 3.26 x 10^-3^), and BCL10 (HR 1.21, 95% CI 1.11-1.31, *q*-val = 4.71 x 10^-3^). RIPPLY1 constitutes a component of a transcriptional repressor complex that has been implicated in acute pancreatitis(*21*). FSTL5 is a secretory protein that has previously been shown to inhibit proliferation and epithelial to mesenchymal transition in HCC cells(*22, 23*). BCL10 is a transcription factor that regulates lymphocyte proliferation(*24*). We also found evidence of an association of CEL, a glycoprotein secreted from the pancreas into the digestive tract, with pancreatic cancer risk (HR 1.30, 95% CI 1.12-1.49, *q*-val = 3.83 x 10^-2^)(*25*).

*Genitourinary cancers*

36 proteins were associated with kidney cancer risk including markers of kidney injury or function (HAVCR1, SPP1), blood pressure regulation (REN), and immune response (CD70, WFDC2)(*26-29*). Notably, after restricting analyses to the first five years of follow-up, there were an additional 57 protein associations not identified in primary analyses including those mapped to B4GALT6 (HR 0.29, 95% CI 0.19-0.44, *q*-val = 6.77 x 10^-6^), CFB (HR 3.01, 95% CI 2.07-4.38, *q*-val = 7.48 x 10^-6)^ and FCGRT (HR 0.40, 95% CI 0.29-0.55, *q*-val = 1.25 x 10^-5)^. Likewise, there were 38 proteins associated with bladder cancer risk, including GTPase NRas, encoded by *NRAS*, a potential oncogene for bladder cancer(*30*). In addition to PSA, 10 proteins were associated with prostate cancer risk including FLT4 (HR 1.26, 95% CI 1.13-1.40, *q*-val = 7.02 x 10^-3^), ANKRD1 (HR 1.18, 95% CI 1.09-1.27, *q*-val = 1.05 x 10^-2^), and ACP3 (HR 1.20, 95% CI 1.10-1.31, *q*-val = 1.33 x 10^-2^). In analyses restricted to the first five years of follow-up, there were an additional 5 protein associations including OAT (HR 1.39, 59% CI 1.21-1.61, *q*-val = 1.60 x 10^-3^) and NCR2 (HR 1.28, 95% CI 1.14-1.44, *q*-val = 4.33 x 10^-3^).

*Gynaecological cancers*

We observed 29 proteins associated with endometrial cancer risk and 39 with ovarian cancer risk. The protein most strongly associated with endometrial cancer risk was DKK4 (HR 1.31, 95% CI 1.17-1.47, *q*-val = 1.27 x 10^-3^), a member of the Dickkopf family which regulates Wnt signalling and has been shown to be upregulated in endometrial tumour tissue as compared to normal and atypical endometrial hyperplasia tissue(*31, 32*). RUNX3 (HR 1.20, 95% CI 1.11-1.30, *q*-val = 2.29 x 10^-3^) has been implicated as a tumour suppressor in endometrial cancer(*33*). 16 additional proteins were associated with endometrial cancer risk when analyses were restricted to the first five years of follow-up including IMPA1 (HR 1.40, 1.25-1.56, *q*-val = 2.90 x 10^-6^), PRPF6 (HR 1.35, 95% CI 1.20-1.51, *q*-val = 1.60 x 10^-4^), and KAT5 (HR 0.38, 95% CI 0.24-0.59, *q*-val = 2.89 x 10^-3^). 15 of these proteins were exclusively associated with endometrial cancer risk within this follow-up period, supporting their potential utlility as specific biomarkers for this disease. After MUC-16 (CA-125), the proteins most strongly associated with ovarian cancer risk included PRND (HR 1.44, 95% CI 1.26-1.64, *q*-val = 8.92 x 10^-5^), CEP76 (HR 1.32, 95% CI 1.18-1.48, *q*-val = 4.72 x 10^-4^), and CDKN2C (HR 1.24, 95% CI 1.13-1.35, *q*-val = 9.36 x 10^-4^), all of which have an unclear link to ovarian cancer, highlighting proteins that merit further investigation in future studies to clarify their relationship with ovarian cancer.

*Upper aerodigestive tract cancers*

131 proteins were associated with risk of squamous cell carcinoma of the upper aerodigestive tract, including PTPRS (HR 0.52, 95% CI 0.42-0.64, *q*-val = 1.16 x 10^-6^), ALDH1A1 (HR 1.58, 95% CI 1.36-1.84, *q*-val = 2.31 x 10^-6^)^,^ and ADAMTSL2 (HR 1.44, 95% CI 1.27-1.62, *q*-val = 8.94 x 10^-6^). PTPRS can dephosphorylate EGFR and is frequently deleted in head and neck squamous cell carcinoma(*34*). ALDH1A1 is a putative cancer stem cell marker that has been shown to promote tumour angiogenesis and metastasis(*35*). 57 proteins were associated with risk of non-squamous cell carcinoma of the upper aerodigestive tract such as HEBP1, CTH, and PANK1, with an additional 20 associated after exclusion of the first five years of follow-up including GFAP(HR 1.44, 95% CI 1.23-1.69, *q*-val = 1.48 x 10^-3^) and TNFRSF14 (HR 1.44, 95% CI 1.23-1.75, *q*-val = 2.75 x 10^-3^).

*Glioma*

We identified 11 proteins associated with glioma risk, most notably RNASE4 (HR 0.79, 95% CI 0.72-0.86, *q*-val = 2.01 x 10^-4^), LRIT2 (HR = 1.27, 95% CI 1.14-1.42, *q*-val = 3.43 x 10^-3^), and RBM18 (HR = 1.33, 95% CI 1.17-1.51, *q*-val = 3.93 x 10^-3^). RNASE4, a member of the ribonuclease A superfamily and potential antimicrobial protein, is a proposed biomarker of prostate cancer prognosis that has an unclear link to glioma risk(*36, 37*).

*Melanoma*

24 proteins were associated with melanoma risk, most notably PRSS35 (HR 1.28, 95% CI 1.18-1.40, *q*-val = 3.06 x 10^-5^), CSNK2A1 (HR 1.32, 95% CI 1.18-1.46, *q*-val = 2.93 x 10^-4^), and ASIP (HR 1.17, 95% CI 1.10-1.26, *q*-val = 2.07 x 10^-3^). ASIP, a secreted protein involved in the regulation of melanogenesis, is a known melanoma susceptibility locus(*38*).

*Thyroid*

22 proteins were associated with thyroid cancer risk with the strongest associations observed for SMR3A (HR 1.25, 95% CI 1.15-1.36, *q*-val = 2.16 x 10^-4^), HMX3 (HR 1.15, 95% CI 1.09-1.23, *q*-val = 8.59 x 10^-4^), NPTX2 (HR 1.43, 95% CI 1.23-1.66, *q*-val = 1.69 x 10^-3^), and TG (HR 1.24, 95% CI 1.12-1.37, *q*-val =6.84 x 10^-3^). HMX3 regulates the production of growth hormone and acromegaly, characterised by excess growth hormone production, has been linked to thyroid cancer risk(*39*). TG, a glycoprotein synthesised by thyroid follicular cells, is used as a marker for residual, recurrent, or metastatic disease in patients with differentiated thyroid cancer(*40*).
