## Supplementary Methods for "Population-scale plasma proteomics reveals pre-diagnostic biomarkers and shared pathways of cancer risk across 25 cancers"

**Supplementary Materials**

**Intra-class correlation coefficients (ICCs) to compare measurements before/after plate correction**

To evaluate the impact of plate correction on assay reproducibility, intra-class correlation coefficients (ICCs) were estimated using the 233 blind duplicate samples available in EPIC. For each aptamer, a linear mixed-effects model was fitted to the combined dataset comprising all EPIC samples and the blind duplicates:

$Y_{j_{r}} =\mu+ \beta_{j} + \zeta_{j_{r}}$

where $Y_{j_{r}}$denotes the r-th measurement of aptamer Y for sample j (r=1 for all EPIC samples, and r=1, …, 233 for the duplciates), $\mu$ is the overall mean of Y, $\beta_{j}\sim N(0, \sigma_{\beta}^{2})$ represents the sample-specific random effect, and $\zeta_{j_{r}}\sim N(0, \sigma_{\zeta}^{2})$ denotes measurement errors and other technical variation.

Under this model, the ICC for aptamer Y was defined as

$$ICC = \frac{Var(\beta_{j})}{[Var(\zeta_{j_{r}}) + Var(\beta_{j})]}$$

which quantifies the proportion of total variance attributable to biological differences between samples. Higher ICC values indicate greater measurement reproducibility.

Figure XX summarizes the distributions of ICC values across the 7,289 aptamers before and after plate correction:

- **Left panel**: box-plots of ICC values before (red) and after (cyan) plate correction
- **Middle panel:** empirical density distributions of ICC values before (red) and after (cyan) plate correction
- **Right panel**: scatter plots comparing ICC values before (x-axis) and after (y-axis) plate correction for all 7,289 aptamers.

Overall, the results indicate that plate correction led to a modest but consistent improvement in measurement reproducibility, as reflected by slightly higher ICC values across most aptamers.

**Covariate selection in EPIC**

Minimally adjusted models across all cancer endpoints were stratified on sex, centre, and age at recruitment. Fully adjusted models across all cancer endpoints were further adjusted for highest level of education attained (none/Primary school completed/Technical or professional school/Secondary school/Longer education/Missing), smoking status and number of cigarettes smoked (7 categories: never, former smoker and < 15 cigarettes/day, former smoker and >15 cigarettes/day, former smoker and number of cigarettes smoked unknown, current smoker and < 15 cigarettes/day, current smoker and >15 cigarettes/day, current smoker and number of cigarettes smoked unknown).

Cancer type-specific analyses were further adjusted for cancer type-specific risk factors as follows:

**Bladder and Kidney cancer**- [cigarette-pack years (5 categories: quintiles, unknown), and BMI (4 categories: <25, 25-29, 30-34, >=35 kg/m2)]

**Pre and post-menopausal** **Breast cancer** - [family history of breast cancer (yes, no or unknown), parity and age at first birth (8 categories: >=3 children <25 years or age not reported, >=3 children 25-29 years, >=3 children >=30 years, 1-2 children < 25 years, 1-2 children 25-29 years or age not reported, 1-2 children >=30 years, number >= 1of children not reported, no children), age menarche (4 categories: <12 years, 12-14 years, 14-16 years, >=16 years), hormone replacement therapy use (2 categories: never/ever), oral contraceptive use (2 categories: never/ever), alcohol intake (6 categories: < 1 g/day, 1-9 g/day, 10-19 g/day, >20 g/day, nondrinkers, unknown), physical activity (6 categories: 60 MET hours per week, unknown), BMI (4 categories: <25, 25-29, 30-34, >=35 kg/m2)

**Colon and rectal cancer** - [family history of colorectal cancer (yes, no or unknown), waist to hip ratio (5 categories: quintiles), processed meat intake (4 categories: quartiles), physical activity (6 categories: quintiles, unknown), BMI (4 categories: <25, 25-29, 30-34, >=35 kg/m2), alcohol intake (5 categories: non drinker, <1g/day, 1-10g/day, 10-20g/day, >=20g/day), and smoking status and years of smoking (never, former <30 years, former >=30 years, former duration unknown, current <30 years, current >=30 years, current duration unknown)]

**Endometrial cancer** [parity and age at first birth (8 categories: >=3 children <25 years or age not reported, >=3 children 25-29 years, >=3 children >=30 years, 1-2 children < 25 years, 1-2 children 25-29 years or age not reported, 1-2 children >=30 years, number >= 1of children not reported, no children), age menarche (4 categories: <12 years, 12-14 years, 14-16 years, >=16 years), hormone replacement therapy use (2 categories: never/ever), oral contraceptive use (2 categories: never/ever), physical activity (6 categories: quintiles, unknown), smoking status and years of smoking (7 categories: never, former <30 years, former >=30 years, former duration unknown, current <30 years, current >=30 years, current duration unknown), BMI (4 categories: <25, 25-29, 30-34, >=35 kg/m2), and menopausal status at blood collection (4 categories: pre-menopause, peri-menopause, post-menopause, surgical post-menopause – bilateral ovarectomy)].

**CLL**, **Multiple Myeloma, Non Hodgkin lymphoma, Glioma** [BMI (4 categories: <25, 25-29, 30-34, >=35 kg/m2)

**Liver cancers, upper aero-digestive cancers, oesophageal cancers** (including subtypes) [BMI (4 categories: <25, 25-29, 30-34, >=35 kg/m2), and alcohol intake (5 categories: non drinker, <1g/day, 1-10g/day, 10-20g/day, >=20g/day)]

**Lung cancer** - [smoking status and years of smoking (7 categories: never, former <30 years, former >=30 years, former duration unknown, current <30 years, current >=30 years, current duration unknown)]

**Ovarian cancer** – [family history of breast cancer (yes, no or unknown), parity and age at first birth (8 categories: >=3 children <25 years or age not reported, >=3 children 25-29 years, >=3 children >=30 years, 1-2 children < 25 years, 1-2 children 25-29 years or age not reported, 1-2 children >=30 years, number >= 1of children not reported, no children), age menarche (4 categories: <12 years, 12-14 years, 14-16 years, >=16 years), hormone replacement therapy use (2 categories: never/ever), oral contraceptive use (2 categories: never/ever), smoking status and years of smoking (7 categories: never, former <30 years, former >=30 years, former duration unknown, current <30 years, current >=30 years, current duration unknown), BMI (4 categories: <25, 25-29, 30-34, >=35 kg/m2), menopausal status at blood collection (4 categories: pre-menopause, peri-menopause, post-menopause, surgical post-menopause – bilateral ovarectomy), and an interaction between BMI (4 categories) and dichotomized menopausal status (2 categories: natural postmenopause or not)]

**Pancreas cancer** - [BMI (4 categories: <25, 25-29, 30-34, >=35 kg/m2), alcohol intake (5 categories: non drinker, <1g/day, 1-10g/day, 10-20g/day, >=20g/day)] and diabetes (yes, no, unknown)]

**Prostate cancer** - [BMI (4 categories: <25, 25-29, 30-34, >=35 kg/m2)]

**Stomach cancer** - [BMI (4 categories: <25, 25-29, 30-34, >=35 kg/m2), and alcohol intake (5 categories: non drinker, <1g/day, 1-10g/day, 10-20g/day, >=20g/day)]

**Thyroid cancer** - [BMI (4 categories: <25, 25-29, 30-34, >=35 kg/m2)]

We did not adjust **melanoma** beyond highest level of education and smoking status and number of cigarettes smoked since there were no known relevant confounders measured in the EPIC data.

**Covariate selection in ARIC**

Minimally adjusted models across all cancer endpoints were adjusted for education level (Visit 1; 3 categories: none/basic education [less than completed high school], intermediate education [completed high school or equivalent], advanced education [at least some college]) and stratified on sex (Visit 1; 2 categories: male, female), race/field center (Visit 1; 5 categories: White/Forsyth County, NC, White/Minneapolis, MN, White/Washington County, MD, Black/Jackson, MS, Black/other field center), and age (Visit 2; 5-year intervals); ID was used as a cluster variable. Fully adjusted models across all cancer endpoints were further adjusted for smoking status and the number of cigarettes smoked (Visits 2 and 1; 7 categories: never, former smoker and < 15 cigarettes/day, former smoker and >15 cigarettes/day, former smoker and number of cigarettes smoked unknown, current smoker and < 15 cigarettes/day, current smoker and >15 cigarettes/day, current smoker and number of cigarettes smoked unknown), except for liver cancer which was only adjusted for smoking status (Visit 2; 3 categories: never, former, current).

Cancer type-specific analyses were further adjusted for cancer type-specific risk factors as follows:

**Bladder and kidney cancer**- [cigarette-pack years (Visit 2; 5 categories: quartiles, unknown), and BMI (Visit 2; 4 categories: <25, 25-<30, 30-<35, >=35 kg/m2)]

**Breast cancer (post-menopausal)** - [family history of breast cancer (Visit 3; 3 categories: yes, no unknown), parity and age at first birth (Visits 1 and 3; 7 categories: >=3 children <25 years or age not reported, >=3 children 25-29 years, >=3 children >=30 years, 1-2 children < 25 years, 1-2 children 25-29 years or age not reported, 1-2 children >=30 years, no children), age menarche (Visit 1; 4 categories: <12 years, 12-13 years, 14-15 years, >=16 years), hormone replacement therapy use (Visit 2; 2 categories: never/ever), oral contraceptive use (Visit 1; 2 categories: never/ever), alcohol intake (Visit 2; 5 categories: < 1 g ethanol intake/day, 1-9 g/day, 10-19 g/day, >20 g/day, nondrinkers), physical activity (Visit 1; 3 indices: work index, sport index, leisure time index), and BMI (Visit 2; 4 categories: <25, 25-<30, 30-<35, >=35 kg/m2)]

**CLL** - **Multiple myeloma – Non-Hodgkin lymphoma** – [BMI (Visit 2; 4 categories: <25, 25-<30, 30-<35, >=35 kg/m2)]

**Colon and rectal cancer** - [waist to hip ratio (Visit 2; 5 categories: quintiles), processed meat intake (Visit 2; 4 categories: quartiles), physical activity (Visit 1; 3 indices: work index, sport index, leisure time index), BMI (Visit 2; 4 categories: <25, 25-<30, 30-<35, >=35 kg/m2), alcohol intake (Visit 2; 5 categories: < 1 g ethanol intake/day, 1-9 g/day, 10-19 g/day, >20 g/day, nondrinkers), and smoking status and years of smoking (Visits 2 and 1; 6 categories: never, former <30 years, former >=30 years, current <30 years, current >=30 years, duration unknown)]

**Endometrial cancer** [parity and age at first birth (Visits 1 and 3; 7 categories: >=3 childrenchildren <25 years or age not reported, >=3 children children25-29 years, >=3 children children>=30 years, 1-2 children children< 25 years, 1-2 children children25-29 years or age not reported, 1-2 children children>=30 years, no children), age menarche (Visit 1; 4 categories: <12 years, 12-13 years, 14-15 years, >=16 years), hormone replacement therapy use (Visit 2; 2 categories: never/ever), oral contraceptive use (Visit 1; 2 categories: never/ever), physical activity (Visit 1; 3 indices: work index, sport index, leisure time index), smoking status and years of smoking (Visits 2 and 1; 6 categories: never, former <30 years, former >=30 years, current <30 years, current >=30 years, duration unknown), BMI (Visit 2; 4 categories: <25, 25-<30, 30-<35, >=35 kg/m2), and menopausal status at blood collection (Visit 2; 5 categories: pre-menopause, peri-menopause, post-menopause (natural), post-menopause (surgical), unknown)]

**Glioma** - [BMI (Visit 2; 4 categories: <25, 25-<30, 30-<35, >=35 kg/m2)]

**Liver and gallbladder cancer** [BMI (Visit 2; 4 categories: <25, 25-<30, 30-<35, >=35 kg/m2), and alcohol intake (Visit 2; 5 categories: < 1 g ethanol intake/day, 1-9 g/day, 10-19 g/day, >20 g/day, nondrinkers)]

**Lung cancer** - [smoking status and years of smoking (Visits 2 and 1; 6 categories: never, former <30 years, former >=30 years, current <30 years, current >=30 years, duration unknown)]

**Ovarian cancer** – [family history of breast cancer (Visit 3; 3 categories: yes, no unknown), parity and age at first birth (Visits 1 and 3; 7 categories: >=3 children <25 years or age not reported, >=3 children 25-29 years, >=3 children >=30 years, 1-2 children < 25 years, 1-2 children 25-29 years or age not reported, 1-2 children >=30 years, no children), age menarche (Visit 1; 4 categories: <12 years, 12-13 years, 14-15 years, >=16 years), hormone replacement therapy use (Visit 2; 2 categories: never/ever), oral contraceptive use (Visit 1; 2 categories: never/ever), smoking status and years of smoking (Visits 2 and 1; 6 categories: never, former <30 years, former >=30 years, current <30 years, current >=30 years, duration unknown), BMI (Visit 2; 4 categories: <25, 25-<30, 30-<35, >=35 kg/m2), menopausal status at blood collection (Visit 2; 5 categories: pre-menopause, peri-menopause, post-menopause (natural), post-menopause (surgical), unknown), and an interaction between BMI (Visit 2; 4 categories) and dichotomized menopausal status (Visit 2; 2 categories: natural postmenopause or not)]

**Pancreas cancer** - [BMI (Visit 2; 4 categories: <25, 25-<30, 30-<35, >=35 kg/m2), alcohol intake (Visit 2; 5 categories: < 1 g ethanol intake/day, 1-9 g/day, 10-19 g/day, >20 g/day, nondrinkers), and diabetes status (Visit 2; 4 categories: normoglycemic, at-risk diabetes, undiagnosed diabetes, diagnosed diabetes)]

**Prostate cancer** - [BMI (Visit 2; 4 categories: <25, 25-<30, 30-<35, >=35 kg/m2)]
